# Genotoxic colibactin mutational signature in colorectal cancer is associated with clinicopathological features, specific genomic alterations and better survival

**DOI:** 10.1101/2023.03.10.23287127

**Authors:** Peter Georgeson, Robert S. Steinfelder, Tabitha A. Harrison, Bernard J. Pope, Syed H. Zaidi, Conghui Qu, Yi Lin, Jihoon E. Joo, Khalid Mahmood, Mark Clendenning, Romy Walker, Elom K Aglago, Sonja I. Berndt, Hermann Brenner, Peter T. Campbell, Yin Cao, Andrew T. Chan, Jenny Chang-Claude, Niki Dimou, Kimberly F. Doheny, David A. Drew, Jane C. Figueiredo, Amy J. French, Steven Gallinger, Marios Giannakis, Graham G. Giles, Ellen L Goode, Stephen B Gruber, Andrea Gsur, Marc J. Gunter, Sophia Harlid, Michael Hoffmeister, Li Hsu, Wen-Yi Huang, Jeroen R Huyghe, JoAnn E. Manson, Victor Moreno, Neil Murphy, Rami Nassir, Christina C. Newton, Jonathan A. Nowak, Mireia Obón-Santacana, Shuji Ogino, Rish K. Pai, Nikos Papadimitrou, John D. Potter, Robert E. Schoen, Mingyang Song, Wei Sun, Amanda E. Toland, Quang M. Trinh, Kostas Tsilidis, Tomotaka Ugai, Caroline Y Um, Finlay A. Macrae, Christophe Rosty, Thomas J. Hudson, Ingrid M. Winship, Amanda I. Phipps, Mark A. Jenkins, Ulrike Peters, Daniel D. Buchanan

## Abstract

**Background and Aims:** The microbiome has long been suspected of a role in colorectal cancer (CRC) tumorigenesis. The mutational signature SBS88 mechanistically links CRC development with the strain of *Escherichia coli* harboring the *pks* island that produces the genotoxin colibactin, but the genomic, pathological and survival characteristics associated with SBS88-positive tumors are unknown.

**Methods:** SBS88-positive CRCs were identified from targeted sequencing data from 5,292 CRCs from 17 studies and tested for their association with clinico-pathological features, oncogenic pathways, genomic characteristics and survival.

**Results:** In total, 7.5% (398/5,292) of the CRCs were SBS88-positive, of which 98.7% (392/398) were microsatellite stable/microsatellite instability low (MSS/MSI-L), compared with 80% (3916/4894) of SBS88 negative tumors (p=1.5×10^-28^). Analysis of MSS/MSI-L CRCs demonstrated that SBS88 positive CRCs were associated with the distal colon (OR=1.84, 95% CI=1.40-2.42, p=1×10^-5^) and rectum (OR=1.90, 95% CI=1.44-2.51, p=6×10^-6^) tumor sites compared with the proximal colon. The top seven recurrent somatic mutations associated with SBS88-positive CRCs demonstrated mutational contexts associated with colibactin-induced DNA damage, the strongest of which was the *APC*:c.835-8A>G mutation (OR=65.5, 95%CI=39.0-110.0, p=3×10^-80^). Large copy number alterations (CNAs) including CNA loss on 14q and gains on 13q, 16q and 20p were significantly enriched in SBS88- positive CRCs. SBS88-positive CRCs were associated with better CRC-specific survival (p=0.007; hazard ratio of 0.69, 95% CI=0.52-0.90) when stratified by age, sex, study, and by stage.

**Conclusion:** SBS88-positivity, a biomarker of colibactin-induced DNA damage, can identify a novel subtype of CRC characterized by recurrent somatic mutations, copy number alterations and better survival. These findings provide new insights for treatment and prevention strategies for this subtype of CRC.

## INTRODUCTION

Globally, colorectal cancer (CRC) is one of the most common and fatal cancers^1^. In the United States, there were an estimated 149,500 new CRC cases and 52,980 deaths attributed to CRC in 2021^2^. CRC is a heterogeneous disease characterized by different molecular subtypes and pathways of tumorigenesis that have clinical implications for prognosis and treatment, as well as importance for understanding disease etiology (Dekker et al., 2019). For example, high-level microsatellite instability (MSI-high)/mismatch repair deficiency (MMRD) occurs in 10%-15% of CRCs and is associated with a favorable prognosis^3^ and response to immune checkpoint inhibition^4^. Somatic mutations in *KRAS*/*NRAS* predict response to anti-EGFR therapy^5^, while the *BRAF* c.1799T>A p.V600E hotspot mutation is used diagnostically to differentiate MSI-high CRC with a sporadic versus inherited (Lynch syndrome) etiology and, when present, implicates tumorigenesis via the serrated pathway. Therefore, molecularly derived subtypes of CRC have diagnostic and clinical utility.

Tumor mutational signatures represent a novel approach to molecular stratification of CRC^6, 7^ as they can characterize tumors by aggregating each observed somatic DNA mutation to present an overall picture of the mutational processes active in the tumor^8^. Consequently, mutational signature profiles can improve our understanding of the etiology underlying individual tumors. The predominant set of mutational signatures published by COSMIC^9^ includes recently added definitions for signatures arising from colibactin-induced DNA damage, namely single base substitution (SBS) signature SBS88 and small insertions and deletions (ID) signature ID18, characterized by single nucleotide variants (SNVs) and short insertions and deletions (indels), respectively, occurring predominantly in T-homopolymer contexts, and thus providing a biomarker of CRC tumorigenesis caused by *pks^+^ E. coli* colibactin-induced DNA damage. Numerous studies have reported a higher prevalence of genotoxic strains of *Escherichia coli* harboring the *pks* island (*pks^+^ E. coli*) in CRC-affected individuals compared with healthy individuals^10–12^ and, more recently, a western-style diet was found to be associated with a higher incidence of CRC containing *pks^+^ E. coli*^1^^3^, further implicating a role in the tumorigenesis of CRC through the production of colibactin^10,1^^4^. Colibactin causes genomic damage in the form of inter-strand cross links^1^^5^ and double-stranded breaks^1^^6^. This specific DNA damage is recognizable via a unique tumor mutational signature originally identified in epithelial organoids exposed to colibactin^1^^7^, normal colorectal epithelial cells^1^^8^ and, more recently, in CRCs^1^^9^, providing a mechanistic link between *pks^+^ E. coli* colibactin-induced DNA damage and CRC etiology. Currently, the clinico-pathological consequences of colibactin-induced DNA damage to the tumor and patient are unknown.

In this study, we investigated the clinico-pathological characteristics, genomic features and CRC-specific survival associated with CRC showing colibactin-induced DNA damage, as measured by SBS88, in a large group of CRC patients that had undergone targeted multi-gene panel sequencing of tumor and matched germline DNA as part of the Genetics and Epidemiology of Colorectal Cancer Consortium (GECCO) and Colon Cancer Family Registry study (CCFR).

## MATERIALS AND METHODS

### Study population

A total of 6,111 tumors were available for the study (**Supplementary Table 1**), consisting of 2,542 CRCs sequenced with a 1.34 megabase (Mb) amplicon targeted panel covering 205 genes^20^ and 3,569 CRCs sequenced with a 1.96Mb capture targeted panel covering 350 genes. Formalin-fixed paraffin-embedded (FFPE) CRC tissue was macrodissected and DNA extracted. Matched germline DNA from either peripheral blood or FFPE normal colonic mucosa was also extracted. Details of the targeted panel sequencing are provided in the supplementary material and details of the study design are provided in **Figure 1**.

**Figure 1:**
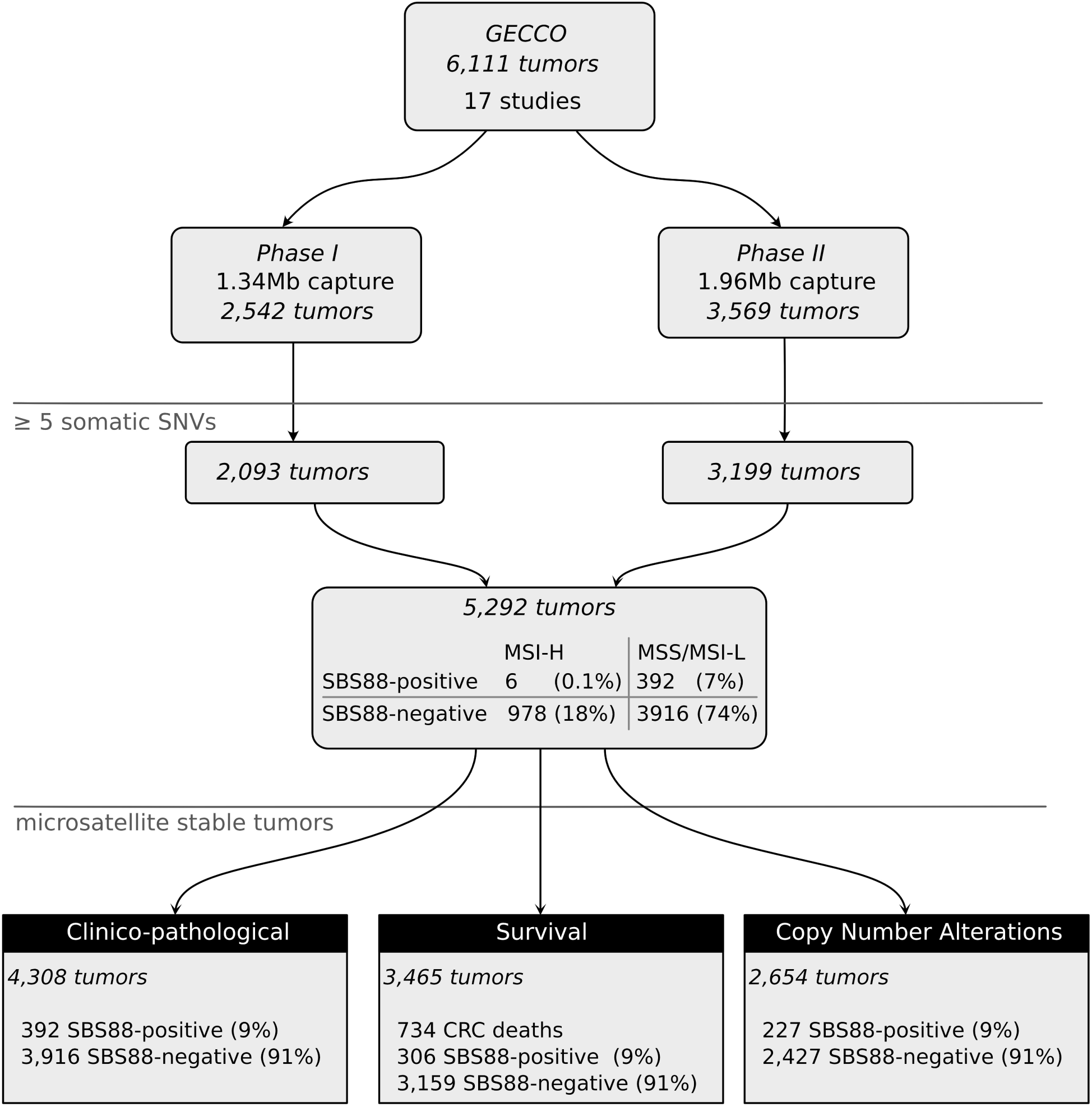
Overview of study design, summarizing included tumors and analyses performed.

### Mutational signatures

Tumor mutational signatures were calculated for each CRC using the simulated annealing method employed by SignatureEstimation^21^. The pre-defined set of 78 COSMIC v3.2 SBS signatures^9^ was reduced to a set of 18 signatures comprising only those previously observed in CRC^22^, including the colibactin-induced signature SBS88; this reduced the potential for mutations to be assigned to signatures less plausible in CRC. This is necessary because targeted panel sequencing produces a limited number of somatic mutations^23^. Only tumors with at least five somatic single nucleotide variants were considered adequate for accurate signature decomposition^24^, leaving 5,292 (86.6%) tumors for further analysis (**Figure 1**). For this study, evidence of *pks^+^ E. coli* colibactin-induced DNA damage was defined as a tumor exhibiting an SBS88 contribution of >10% (SBS88-positive CRC). For indel mutations, 84% of the tumors had a count of <5, meaning these tumors would not be suitable for accurate ID signature decomposition^24^, and, therefore, ID signatures were not studied in this analysis. Similarly, doublet signatures^22^ were not analyzed given their low prevalence in CRC^22^.

### Data collection

The clinico-pathological features assessed included sex (male/female), self-reported race and ethnicity (American Indian or Alaska Native/Asian/Black or African American/White/Other), age at CRC diagnosis (years), tumor site (ICD-9 codes 153.0, 153.1,153.4, 153.6=Proximal/153.2,153.3, 153.7=Distal/154.0, 154.1=Rectal), AJCC tumor stage (1, 2, 3, 4), family history of CRC (yes/no for having one or more first-degree relatives with CRC), and inflammatory bowel disease (yes/no). Information on CRC-specific survival was available from cancer registries, vital statistics registries and study follow up, as described previously^25^.

### Bioinformatics analyses

The genomic characteristics of the CRCs assessed included microsatellite instability status (defined as MSI-high or MSS/MSI-L) determined by mSINGS^26^ and MSIsensor2^27^ and described in further detail by Zaidi et al^20^; tumor mutational burden (TMB; number of somatic SNVs and indels per Mb sequenced); non-silent mutations in known CRC driver genes^28^ (*APC, KRAS, TP53, PIK3CA, PTEN, POLE, CTNNB1, RNF43, SMAD4*) including mutational hotspots e.g. *BRAF*^V600E^ and *KRAS* codon 12/13; tumorigenesis pathways (as described in Zaidi et al^20^) and any recurrent somatic mutations observed in the dataset. Variant allele fraction (VAF) was used to estimate clonal versus sub-clonal mutations for the recurrent somatic mutations identified to be associated with SBS88-positive CRC, with high VAFs suggesting clonality and consequently likely to be an earlier event, rather than the consequences of tumorigenesis^29^. Differences in VAF distributions were assessed by applying a t-test to the VAFs observed for the recurrent mutations relative to the VAFs observed in other mutations from the same tumors.

Somatic copy number alterations (CNAs) were calculated on a subset of 2,654 tumors tested with the 1.96Mb targeted panel that used hybridization capture-based sequencing (compared with amplicon-based sequencing used for the 1.3Mb targeted panel) (**Figure 1**). Three types of CNAs were identified: 1) Focal CNAs (gene-level) were calculated with the GATK 4.1 copy number calling pipeline to identify gene specific CNAs; 2) medium-sized CNAs were calculated by merging gene-level CNA calls into contiguous 10Mb segments^30^ and, to reduce the incidence of false positives, a set of high-confidence CNA calls was generated by considering only segments containing three or more gene-level CNAs indicating the same copy number gain or loss; and 3) large CNAs affecting whole chromosome arms were identified as the most prevalent CNA call across the entire arm. All chromosomes were considered in the analysis except chromosome X and chromosome Y which had insufficient coverage and sex-related bias. P-values were calculated by comparing the proportions of loss or gain in the SBS88 positive CRCs to the proportion observed in the SBS88 negative CRCs in each 10Mb segment, each segment consisting of at least three concordant CNA calls, with Fisher’s exact test.

Stratification of SBS88 positive tumors was explored by performing unsupervised clustering of genomic features. Dimensionality reduction was performed by applying multiple correspondence analysis (MCA)^31^ to features that were significantly enriched in the SBS88 positive tumors compared to SBS88 negative tumors and relatively frequent (>10%). The optimal number of clusters was determined using the “elbow” method applied to the inertia^32^, silhouette^33^ and gap statistic^34^ from each clustering (considering k from 1 to 15), then clusters were assigned using the k-means clustering algorithm^35^.

We used Cox proportional hazards regression to estimate hazard ratios for the association of SBS88 with CRC-specific survival and stratified by sex, study (13/17 studies with available data), stage and dichotomized age of diagnosis (below/above 50 years). Samples with greater than 5-year survival (1,825 days) since recruitment were right-censored. The proportionality assumption held for all analyses.

### Statistical analyses

All statistical analyses were performed using the *scipy* statistical package^36^ on Python 3.7. Unless otherwise stated, two-sided unpaired t-tests were used to calculate p-values when comparing means of two groups, Fisher’s exact test was used to calculate p-values when counting across two groups, Chi-square was used to calculate p-values when comparing counts across more than two groups, and ANOVA was used to calculate p-values for means across more than two groups.

Statistical significance was considered to be p<0.05. For the recurrent somatic mutation analysis, adjusted p-values were calculated with the Benjamini-Hochberg procedure^37^ with a false discovery rate of 0.05.

## RESULTS

In total, 398 of the 5,292 included tumors (7.5%) across 17 studies demonstrated the presence of the SBS88 signature, with the proportion of SBS88 observed in each tumor ranging from 10.1% to 72.5% (mean ± standard deviation (SD) = 21.1 ± 10.5%) (**Figure 1**; **Supplementary Table 1**). Of these SBS88 positive CRCs, 98.7% (392/398) were microsatellite stable / microsatellite instability low (MSS/MSI-L), compared with 80% (3916/4894) of SBS88 negative tumors (p=1.5×10^-28^; Fisher’s exact test) (**Supplementary Figure 1**). Due to the predominance of the SBS88 positive CRCs in the MSS/MSI-L subtype and the heterogeneity and biology introduced by MSI-high, subsequent analyses focused only on MSS/MSI-L tumors.

### SBS88-positive tumors are associated with specific clinico-pathological features

Of the clinico-pathological features assessed, age at CRC diagnosis, sex, and tumor site demonstrated an association with SBS88. SBS88 positive CRCs were more common in the distal colon (OR=1.84, 95% CI=1.40-2.42, p=1×10^-5^; Fisher exact test) and rectum (OR=1.90, 95% CI=1.44-2.51, p=6×10^-6^) compared with the proximal colon (**Table 1**). Further stratification by tumor site demonstrated the highest proportion of SBS88 positive CRCs in the sigmoid, rectosigmoid junction, and rectum (**Supplementary Figure 2**). Female sex (57.7% vs 50.7%; p=0.01) and younger mean age at CRC diagnosis (64.6 ± 11.9 vs 66.0 ± 11.7; p=0.03) were associated with SBS88 positive CRC (**Table 1**; **Supplementary Figure 3**).

**Table 1:**
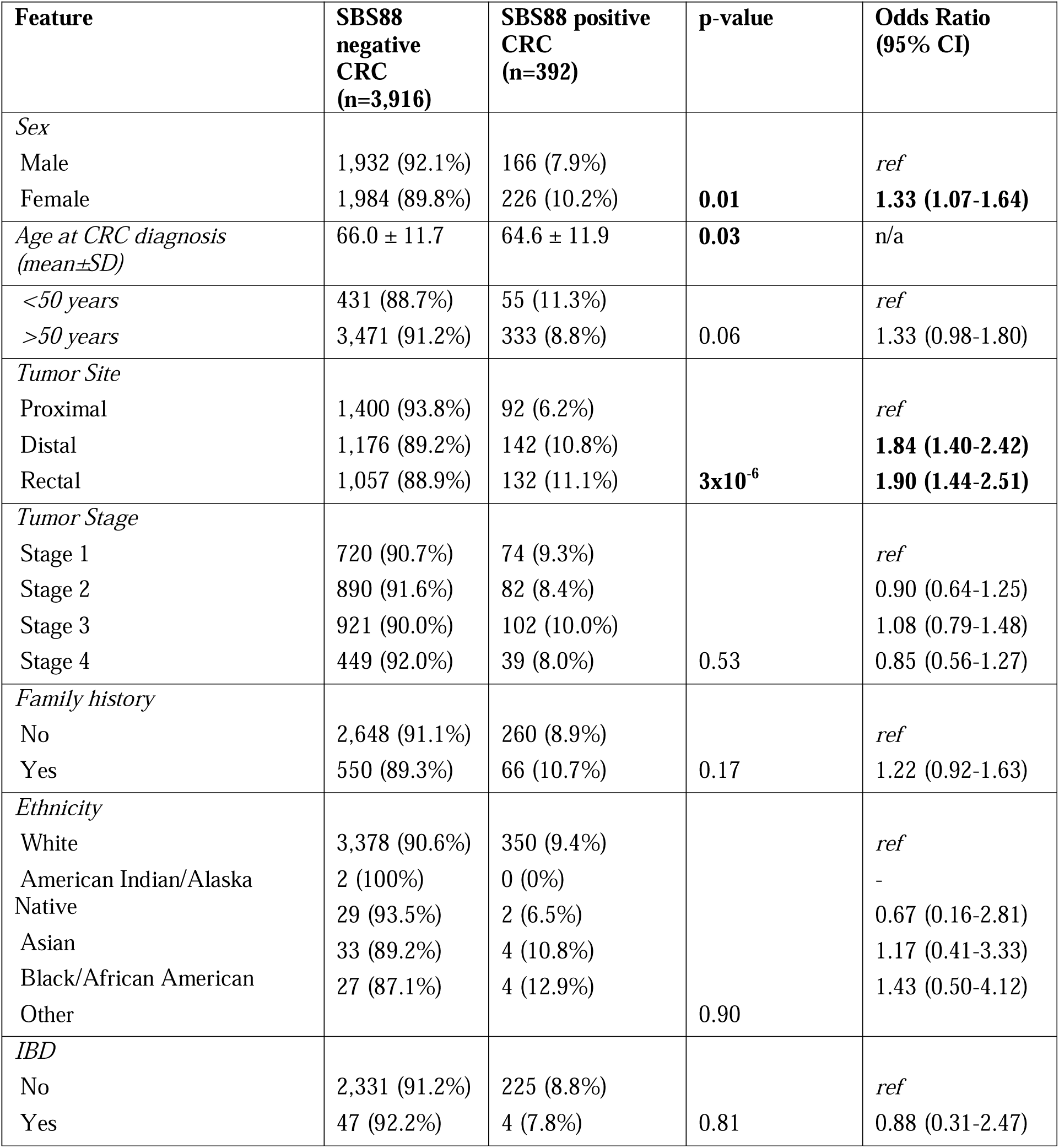
Comparison of clinico-pathological features across MSS/MSI-L CRC (n=4,308) stratified by SBS88 negative and SBS88 positive CRCs. P-values were either calculated from a Chi-square one-way test, or a two-sided unpaired t-test (age of diagnosis). Statistically significant p-values (<0.05) are highlighted in bold.

### SBS88-positive tumors are associated with unique genomic characteristics

The mean tumor mutational burden (TMB) was 6.3 ± 3.2 mutations/Mb in the SBS88 positive CRCs which was significantly lower than in SBS88 negative CRCs (11.1 ± 31.6 mutations/Mb; p=3×10^-3^) (**Table 2**). Tumors with *POLD1* or *POLE* non-synonymous exonuclease domain mutations were observed only in the SBS88-negative tumors (23 and 61 (0.6% and 1.6%) out of 3,916, respectively) (**Table 2)**; the difference in mean TMB remained significant with the *POLE*/*POLD1* tumors excluded (8.3 ±17.7 mutations/Mb; p=0.03). Several somatic CRC driver genes and oncogenic pathways were negatively associated with SBS88 positive CRC (**Supplementary Table 2**). When somatic hotspots were considered, there were 24 recurrent somatic mutations significantly associated with SBS88 status (p<0.05) that were seen in at least three SBS88 positive or SBS88 negative tumors including in *APC*, *SMAD4*, *TP53*, *PIK3CA*, *KRAS* and *BRAF* genes, where 18 mutations were positively associated (**Figure 2a**) and six negatively associated (**Supplementary Figure 4**), demonstrating a high degree of mutual exclusivity. *APC* c.835-8A>G was the strongest recurrent mutation associated with SBS88 positive CRC (OR=65.5, 95%CI=39.0-110.0, p=3×10^-80^) (**Table 2**, **Figure 2a)**. The top seven recurrent somatic mutations associated with SBS88-positive CRCs ranked by lowest p-value (all with adjusted p<0.01) demonstrated mutational contexts associated with colibactin-induced DNA damage (ATT>C, TTT>A, and ATA>C) and included *APC*:c.835-8A>G (ATT>C), *SMAD4*:c.788-8A>G (ATT>C), *APC*:c.1549-8A>G (ATT>C), *APC*:c.1600A>T (p.Lys534Ter) (TTT>A), *TP53*:c.659A>G (p.Tyr220Cys) (ATA>C), *PIK3CA*:c.3127A>G (p.Met1043Val) (ATT>C) and *IWS1*:c.872A>G (p.Asn291Ser) (ATT>C) (**Table 2**, **Figure 2a**). The proportion of tumors with these top seven recurrent somatic mutations by the SBS88 proportion is shown in **Figure 2b**, where the *APC* c.835-8A>G hotspot mutation, the most prevalent recurrent somatic mutation associated with SBS88-positive CRCs, demonstrated increased prevalence in tumors with higher levels of SBS88.

**Table 2:** Comparison of genomic features and recurrent somatic mutations across MSS/MSI-L CRC (n=4,308) stratified by SBS88 negative and SBS88 positive CRCs. P-values were either calculated with Fisher exact (2×2 categorical) Chi-square one-way test (>2 categorical), or from a two-sided unpaired t-test (TMB, CNA count). P-value and odds ratios for copy number alterations are for the least significant segment in each contiguous region. Statistically significant p-values (<0.05) are highlighted in bold. Only recurrent somatic mutations with p<0.0001 are included, all of which remain significant after Benjamini-Hochberg adjustment (FDR<0.05). ^†^adj=Benjamini-Hochberg adjusted p-value.

| Feature | SBS88 negative CRC (n=3,916) | SBS88 positive CRC (n=392) | p-value | Adj<sup>†</sup> p-value | Odds Ratio (95% CI) |
| --- | --- | --- | --- | --- | --- |
| *TMB (mutations/Mb)* | 11.1 ± 31.6 | 6.3 ± 3.2 | **3x10<sup>-3</sup>** |  | n/a |
| *CNA count* | 365.0 ± 191.7 (n=2,427) | 378.9 ± 186.0 (n=227) | 0.29 |  | n/a |
| *POLE exo* | 61 (100%) | 0 (0%) | **0.01** |  | - |
| *POLD1 exo* | 23 (100%) | 0 (0%) | 0.13 |  |  |
| *No POLE/POLD1 exo* | 3,837 (90.7%) | 392 (9.3%) |  |  |  |
| *Recurrent somatic mutations with SBS88 related contexts* |  |  |  |  |  |
| *APC:c.835-8A>G* |  |  |  |  |  |
| Absent | 3,898 (92.8%) | 301 (7.2%) |  |  |  |
| Present | 18 (16.5%) | 91 (83.5%) | **3x10<sup>-80</sup>** | **2x10<sup>-78</sup>** | **65.5 (39.0-110.0)** |
| *APC:c.1549-8A>G* |  |  |  |  |  |
| Absent | 3,915 (91.1%) | 383 (8.9%) |  |  |  |
| Present | 1 (10%) | 9 (90%) | **4x10<sup>-9</sup>** | **2x10<sup>-7</sup>** | **92.0 (11.6-728.1)** |
| *SMAD4:c.788-8A>G* |  |  |  |  |  |
| Absent | 3,915 (91.0%) | 386 (9.0%) |  |  |  |
| Present | 1 (14.6%) | 6 (85.7%) | **4x10<sup>-6</sup>** | **1x10<sup>-4</sup>** | **60.9 (7.3-506.8)** |
| *APC:c.1600A>T (p.Lys534Ter)* |  |  |  |  |  |
| Absent | 3,913 (91.0%) | 385 (9.0%) |  |  |  |
| Present | 3 (30%) | 7 (70%) | **5x10<sup>-6</sup>** | **1x10<sup>-4</sup>** | **23.7 (6.1-92.1)** |
| *TP53:c.659A>G (p.Tyr220Cys)* |  |  |  |  |  |
| Absent | 3,897 (91.1%) | 381 (8.9%) |  |  |  |
| Present | 19 (63.3%) | 11 (36.7%) | **3x10<sup>-5</sup>** | **6x10<sup>-4</sup>** | **5.9 (2.8-12.6)** |
| *PIK3CA:c.3127A>G (p.Met1043Val)* |  |  |  |  |  |
| Absent | 3,916 (91.0%) | 388 (9.0%) |  |  |  |
| Present | 0 | 4 (100%) | **7x10<sup>-5</sup>** | **1x10<sup>-3</sup>** | - |
| *IWS1:c.872A>G p.Asn291Ser* |  |  |  |  |  |
| Absent | 3,915 (91.0%) | 388 (9.0%) |  |  |  |
| Present | 1 (20.0%) | 4 (80.0%) | **3x10<sup>-4</sup>** | **0.004** | **40.36 (4.5-362.0)** |

**Figure 2:**
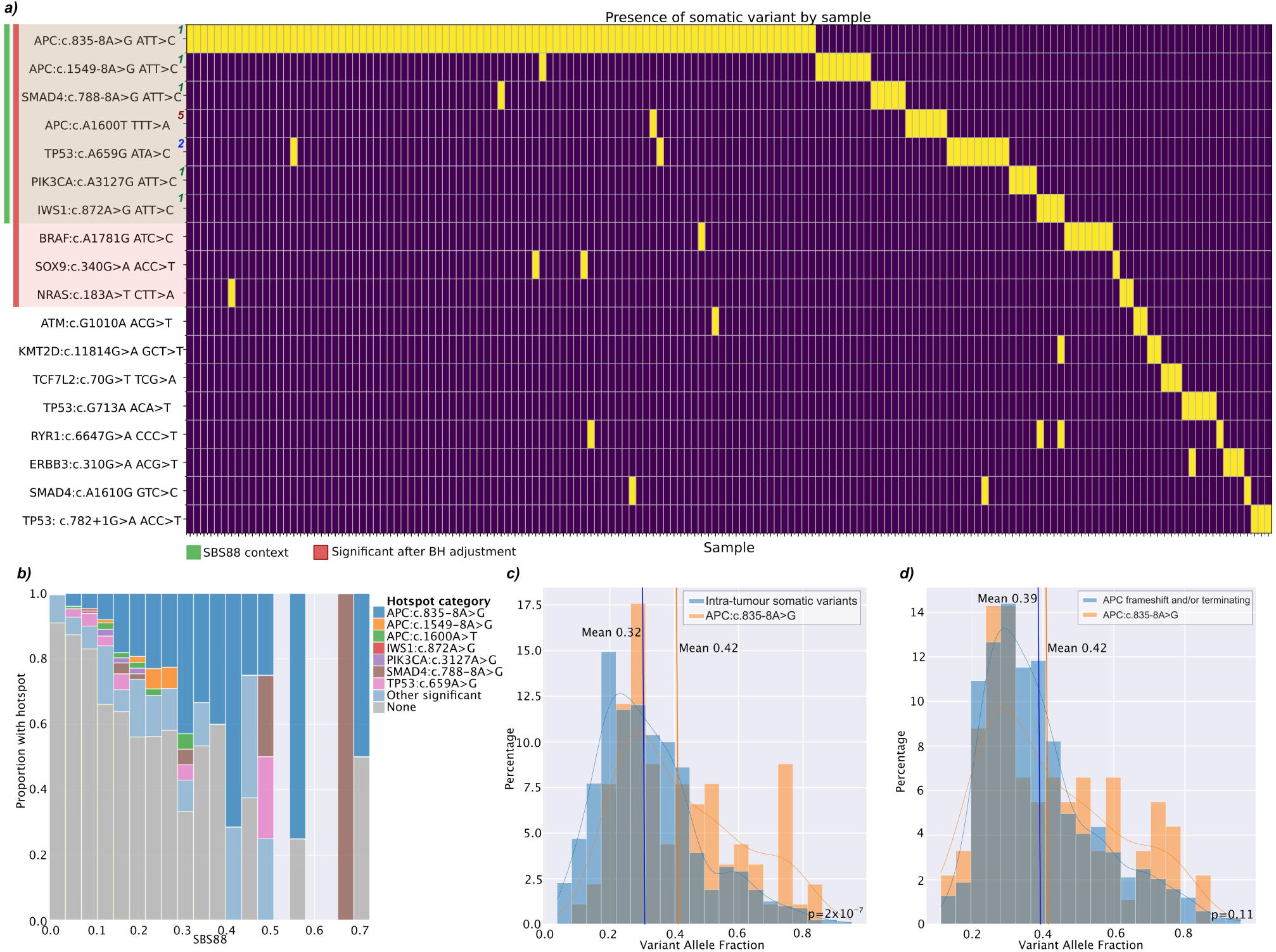
**(a)** Somatic mutations (n=18) seen in at least three SBS88 positive CRCs that were significantly enriched (p<0.05) compared with SBS88 negative CRCs. Somatic mutations are ranked by p-value with the seven most significant mutations located in genomic contexts that are major contributors to the SBS88 signature (context rank in SBS88 shown as superscript). The *APC*, *SMAD4* and *TP53* genes had more than one recurring somatic mutation associated with SBS88 positive CRC. Ten mutations remained significant after Benjamini-Hochberg adjustment (FDR 0.05). **(b)** The positive relationship between the increasing proportion of the SBS88 mutational signature and the likelihood of observing a significant mutational hotspot, in particular *APC*:c.835-8A>G. The seven significant recurrent mutations in an SBS88 context are included individually, as well as the 11 other positively associated recurrent mutations. **(c, d)** Variant allele fraction of the *APC*:c.835-8A>G somatic hotspot mutation in 91 SBS88 positive CRCs is (**c**) significantly higher than other somatic mutations observed in the same tumors, suggesting the hotspot mutation is likely to be clonal and a relatively early somatic event, and (**d**) similar to other truncating and/or frameshifted *APC* mutations, suggesting the hotspot mutation is a likely driver mutation akin to these pathogenic *APC* mutations.

### The SBS88-associated recurrent mutation APC:c.835-8A>G is a driver of CRC tumorigenesis

We assessed the potential for recurrent mutations to constitute driver events by inferring clonality. The recurrent variants *APC*:c.835-8A>G and *TP53*:c.659A>G (p.Tyr220Cys) observed in SBS88-positive tumors showed a significantly higher variant allele fraction (VAF) than the other somatic variants observed in these tumors with mean±SD 0.42±0.19 vs 0.32±0.16 (p=2×10^-7^; t-test) and 0.54±0.19 vs 0.31±0.18 (p=2×10^-4^), respectively, with the mean higher VAFs for these mutations suggesting that they are clonal and therefore an earlier somatic event (**Figure 2c**). The mean VAFs for the *APC*:c.835-8A>G mutation (0.42±0.19) were consistent with the mean VAFs for 1,327 frameshift and truncating *APC* somatic mutations observed in 1,207 tumors (0.39±0.16) supporting the *APC*:c.835-8A>G mutation as a driver mutation (**Figure 2d**).

Stratified analyses performed on the SBS88 positive CRCs by the presence or absence of the *APC*:c.835-8A>G mutation demonstrated a significant enrichment of the sigmoid and rectosigmoid tumor site for SBS88 positive CRCs with the *APC*:c.835-8A>G mutation (p=8×10^-9^; **Supplementary Table 3**). SBS88 positive CRCs with the *APC:*c.835-8A>G mutation had higher proportions of SBS88 signature compared with SBS88 positive CRCs without the *APC:*c.835-8A>G mutation (mean ± SD: 27.8 ± 12.9%, n=91 vs 19.1 ± 8.7%, n=301; p=6×10^-8^) (**Figure 2b**).

### SBS88-positive tumors have a genomic profile that includes large-scale copy-number alteration events

Copy number alterations (CNA) analysis was performed at gene (focal), medium (10Mb), and chromosome arm level on 2,654 tumors that used hybridization capture-based sequencing (**Figure 1**, **Supplementary Table 4**). At gene level, the median number of CNA gains or CNA losses in SBS88-positive CRCs compared with the SBS88 negative CRCs is shown in **Supplementary Table 5**. There were 226 significant (p<0.05) (115 enriched in SBS88 positive, 111 enriched in SBS88 negative) gene level CNA events detected when comparing the 227 SBS88 positive CRCs to 2,427 SBS88 negative CRCs (**Figure 3**), with each significant CNA on average affecting 62 (27%) of SBS88 positive CRCs. For the subset of 49 SBS88 positive CRCs with the *APC* c.835-8A>G hotspot mutation with CNA data, 289 significant gene level CNAs were identified when compared with 2,368 SBS88 negative tumors without *APC* c.835-8A>G (**Supplementary Figure 5a**). In contrast, only 29 significant gene level CNA events were observed in 174 SBS88 positive CRCs without *APC* c.825-8A>G when compared with the SBS88 negative tumors (**Supplementary Figure 5b**). A high proportion of the gene level CNAs were observed to cluster proximally on the genome – for example, 39 of the most significant 44 enriched CNA events are losses on chromosome 14 with the remaining five significantly enriched CNAs being losses on chromosome 2 suggesting the presence of larger copy number events.

**Figure 3:**
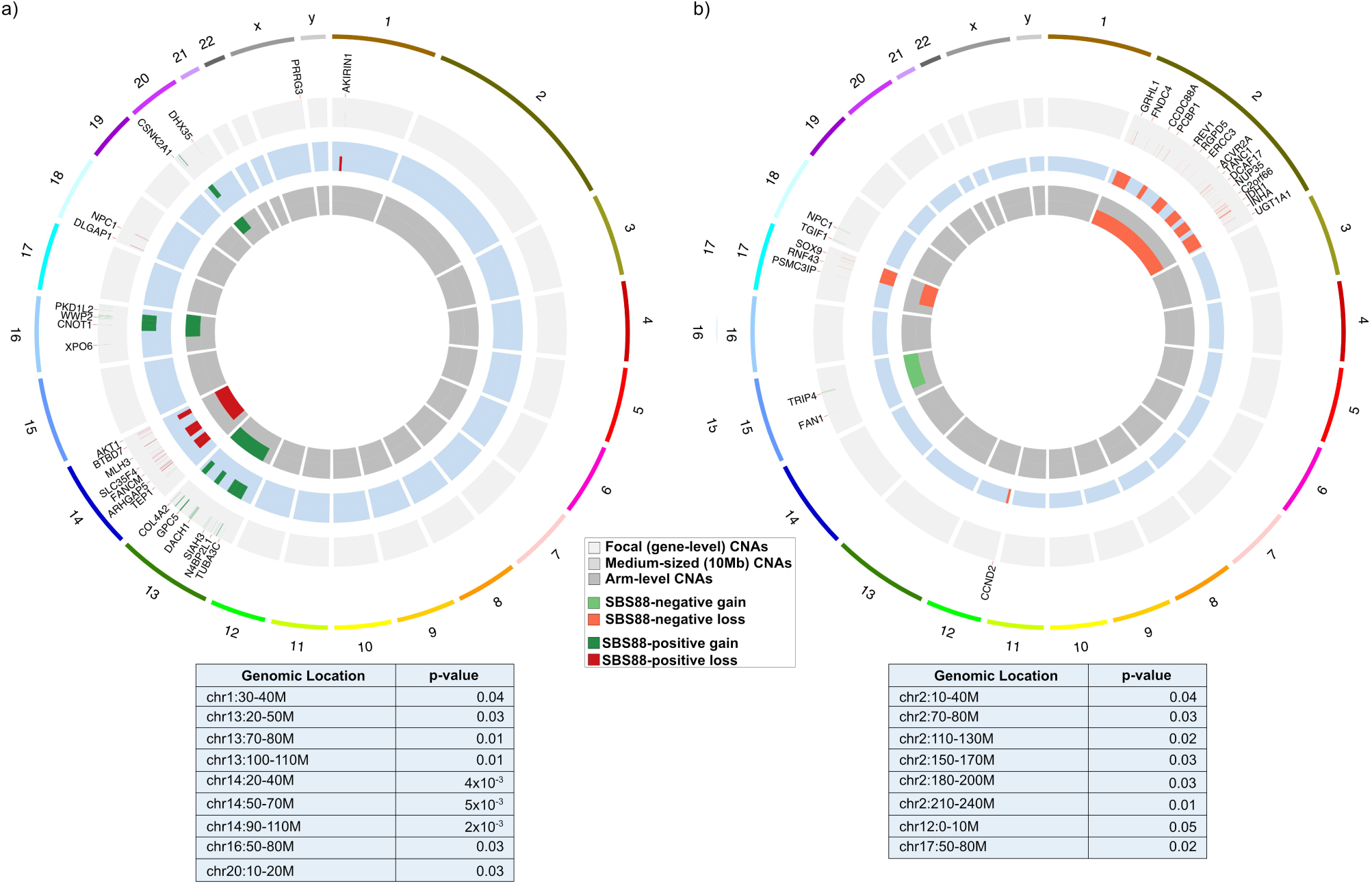
Circos plots showing the overlap between gene (focal) (n=226), medium (10Mb) (n=17) and chromosome arm (n=7) level copy number alterations (CNAs) (losses in red, gains in green), that were significantly enriched in either (a) SBS88-positive tumors or (b) SBS-negative tumors. The outer ring shows gene-level (focal) events, with medium (10Mb) and chromosome arm-level events on the middle and inner rings, respectively. P-values of significant medium sized CNAs are listed below each circos plot.

For medium sized CNAs, gene level CNAs were merged into contiguous 10Mb segments^30^ and a high-confidence set obtained by only considering segments containing at least three genes with the same CNA change (i.e. all gene-level events within a segment being gains or all being losses). There were 33 unique 10MB segments observed to be significantly different between the SBS88 positive and negative CRCs, forming 17 contiguous regions, including copy number losses on chromosomes 1, 2, 12, 14 and 17, and gains on chromosomes 13, 16 **and 20 (**Supplementary Table 5, Supplementary Figure 6 **and** Supplementary Figure 7).

Further clustering of medium CNAs into chromosome arm level CNAs demonstrated a CNA loss on 14q and gains on 13q, 16q and 20p that were significantly enriched in SBS88 positive CRCs and CNA losses on chromosome 2p, 2q, and 17q and a gain on chromosome 15q that were significantly underrepresented in SBS88 positive CRCs (**Figure 3** and **Supplementary Table 6**). CNA events significantly enriched or underrepresented in SBS88 positive tumors with and without *APC* c.835-8A>G are shown in **Supplementary Figures 5a-d**.

We observed associations between SBS88, TMB and CNA counts, with SBS88 levels showing a significant inverse relationship to TMB (p=0.003; excluding 79 tumors with *POLE/POLD1* exonuclease mutations: p=0.03) and non-significant positive relationship to CNA count (p=0.06) (**Supplementary Figures 8a & 8b**, respectively). Individual hotspots also exhibited different levels of SBS88 (p=8×10^-6^; **Supplementary Figure 8d**) and TMB (p=4×10^-6^; **Supplementary Figure 8e**), but not CNA count (p=0.41; **Supplementary Figure 8f**), with *APC*:c.835-8A>G and *SMAD4*:c.788-8A>G showing significantly elevated SBS88 proportions (p=4×10^-7^ and p=0.01, respectively; **Supplementary Figure 8d**), and *TP53*:c.659A>G, *SMAD4*:c.788-8A>G, *PIK3CA*:c.3127A>G and *IWS1*:c.872A>G showing elevated TMB (p=2×10^-4^, p=0.04, p=3×10^-22^ and p=3×10^-18^, respectively; **Supplementary Figure 8e**) compared with other SBS88-positive tumors.

### SBS88-positive tumors cluster into groups exhibiting distinct genomic characteristics

The observation that specific genomic and tumor features that were significantly associated with SBS88-positive CRCs were correlated (**Supplementary Figure 9**) suggested the likelihood of molecular heterogeneity in SBS88 positive CRCs. Clustering SBS88 positive tumors based on enriched genomic features with sufficient frequency (>10%) revealed the presence of three clusters (**Figure 4, Supplementary Figure 10)**. ***Cluster 1*** (*APC* hotspot/20p gain cluster) was the largest cluster comprising predominantly distal and rectal tumors and characterized genomically by both the *APC* c.835-8 A>G hotspot and gains on the 20p chromosomal arm. ***Cluster 2*** (*TP53*/CNA dominant cluster) showed pathogenic *TP53* mutations and the highest proportion of SBS88 associated CNAs, namely 16q gain, 13q gain, and 14q loss with a predominance for distal and rectal tumors. ***Cluster 3*** (WNT/CNA rare cluster) tumors were absent of the SBS88 associated large CNAs but instead harbored *KRAS* and *TGF*β mutations, with >90% of tumors in this cluster demonstrating WNT pathway activating mutations, with relatively more of these tumors in the proximal colon. A summary of each cluster, its characteristics and associated features are shown in **Supplementary Tables 7** and **8**. The molecular profile of SBS88 positive CRC by these three clusters is demonstrated in **Supplementary Figure 11**.

**Figure 4:**
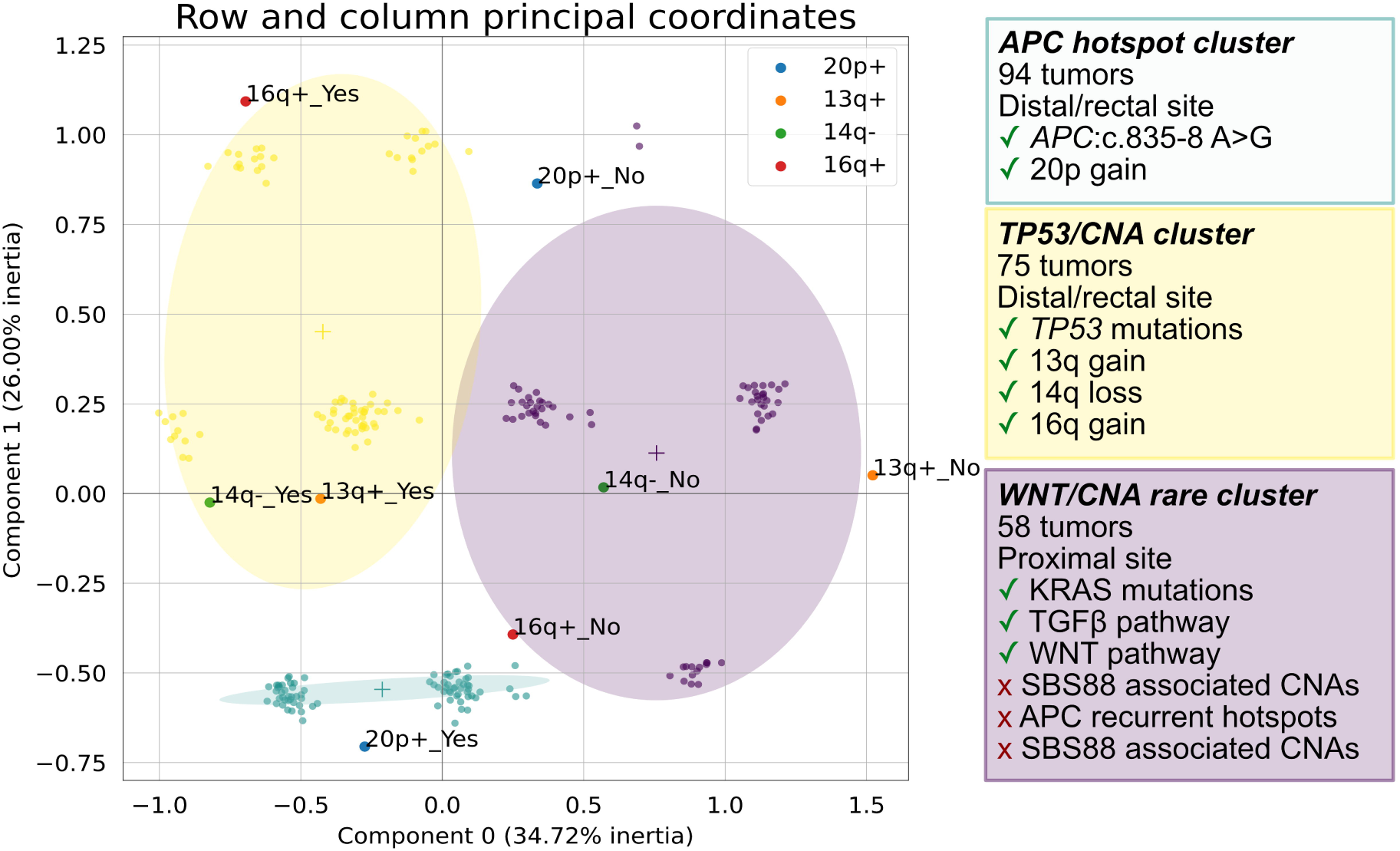
clustering SBS88 positive tumors on enriched and relatively frequent genomic features revealed potential heterogeneity. Each unlabeled point represents a tumor in 2D MCA (multiple correspondence analysis) space, with color representing cluster membership (green=cluster 1; yellow=cluster 2; purple=cluster 3). Ellipses show the 95% confidence interval for cluster membership. Labeled points show genomic features mapped to the 2D MCA space, with proximity of features representing association between features.

### SBS88-positive tumors are associated with better CRC-specific survival

We assessed CRC-specific survival between the SBS88 positive and SBS88 negative CRCs, in the subset of MSS/MSI-L tumors with available data (n=3,465; **Figure 1; Supplementary Table 1**). This subset of tumors included 734 CRC-related deaths. Although CRC-specific survival between the groups was not significantly different (p=0.06; **Figure 5**), after stratifying by age, sex, study, and by stage, the SBS88 positive CRCs were associated with better survival (p=0.007) with a hazard ratio of 0.69 (95% CI 0.52 to 0.90). Tumor site was not significant with this set of included features (**Supplementary Figure 12**). When stratified analyses were performed on the subset of SBS88 positive CRCs with the *APC* c.835-8A>G hotspot mutation, a similar improved survival effect was observed although it was no longer significant (p=0.21). When considering significant CNA chromosome arm changes between SBS88 positive tumors to SBS88 negative tumors, gains on chromosome 15q and losses on 17q were associated with poorer survival across the cohort (p=0.002, OR=1.33-3.44, and 0.04, 1.01-1.61, respectively) (**Supplementary Table 6**). Survival analysis on SBS88-positive tumors with both CNA and survival data (n=168) comparing the three clusters did not reveal a significant difference in survival (p=0.4), though a trend of improved survival in cluster 3 (WNT/CNA rare) relative to the other two clusters was observed (**Supplementary Figure 13**).

**Figure 5:**
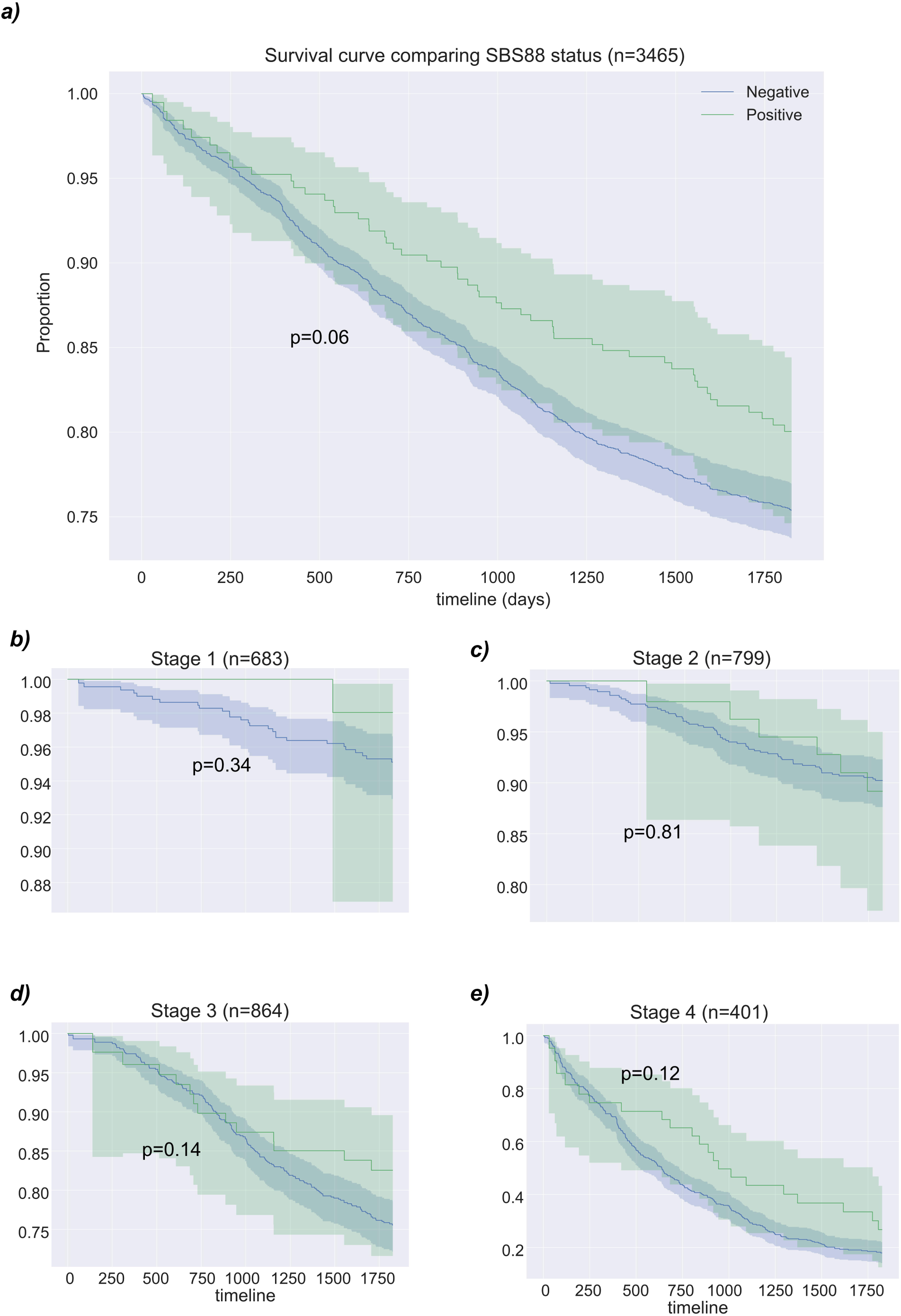
Kaplan-Meier survival curve with CRC-specific death, showing 95% confidence interval (unadjusted). The log-rank comparison of survival curves is not statistically significant across the cohort (p=0.06) **(a)**, or for individual stages **(b-e)** but with stratification on study, sex, stage and dichotomized age the Cox proportional hazards model finds presence of SBS88 as significant (p=0.007).

## DISCUSSION

In this large study of genomically characterized CRCs, we describe a novel subtype of CRC characterized by the SBS88 tumor mutational signature. These tumors are predominantly MSS/MSI-L and, compared with MSS/MSI-L CRC tumors without this signature, are more likely to occur in the distal colon and rectum, driven by the *APC* c.835-8A>G recurrent hotspot mutation, among other recurrent mutations matching the genomic contexts associated with SBS88, and displaying associations with copy number loss on chromosome 14q, and copy number gains on chromosomes 13q, 16q and 20p. These SBS88-associated genomic features revealed three molecular subtypes of SBS88 positive CRC. Furthermore, SBS88 positive CRCs were associated with better CRC survival compared with SBS88 negative CRCs when stratified on sex, stage, age and study.

The *APC* c.835-8A>G somatic variant was highly enriched in the SBS88 positive CRCs, and importantly, rarely arises in tumors not exhibiting the SBS88 signature, suggesting the variant may be associated with DNA damage induced by colibactin. The genomic context surrounding this variant is consistent with the 3bp context enriched in the SBS88 signature and has previously been associated with colibactin damage^17, 19^. A link between the *APC:*c.835-8A>G mutation and colibactin damage was proposed in a smaller study, although the relationship with the SBS88 signature was not investigated^38^. In addition, our analysis of variant allele fraction distributions suggested this variant is likely clonal and, therefore, an early somatic event, consistent with current proposals that SBS88 is likely the result of early life exposure to colibactin^18, 39, 40^. Combined with its location within a known CRC driver gene, this provides further evidence of its potential status as a driver mutation and the likely importance of the genotoxic colibactin DNA damage targeting this hotspot DNA sequence in the *APC* gene for tumor development. The presence of the *APC* c.835-8A>G mutation and SBS88 in pre-malignant adenomas would support this concept.

Other associated recurrent mutations exhibited genomic contexts reflecting the SBS88 signature definition: five of the top seven associated mutations match the ATT>C context. These SBS88 associated somatic mutations were largely mutually exclusive. The strong association between these somatic variants and SBS88 positive CRC and their rarity in SBS88 negative CRC indicates these specific variants may serve as biomarkers or proxies for the SBS88 mutational signature, which may be of particular importance for identifying colibactin-induced CRC at lower somatic mutation counts where tumor mutational signatures become less reliable due to an increase in reconstruction error^6, 24^.

The specific CNA events associated with SBS88 positive CRC may relate to the mechanism by which colibactin induces DNA damage. Colibactin-induced DNA damage manifests genomically as interstrand crosslinks^15^ and double stranded breaks^19, 41^, which may explain the association with specific CNA events observed in SBS88 positive tumors. Therefore, colibactin may directly cause double stranded breaks at specific genomic sequence/contexts (e.g. T-homopolymers) leading to CNAs. Alternatively, these recurrent CNA events may be an indirect consequence of colibactin-induced DNA damage resulting from increased genomic instability caused by mutations in *TP53* gene or activation of the interstrand crosslink repair mechanism mediated by the Fanconi-anemia pathway, which as a side-effect tends to create double stranded breaks^42^. We observed a cluster of SBS88 positive CRCs characterized by *TP53* mutations and high CNA load (cluster 2). The timing of when these SBS88 associated CNAs occur during the tumorigenesis process may help to resolve this mechanism. We hypothesize that the enrichment of specific CNAs in SBS88 positive CRC may help drive tumorigenesis in a low somatic SNV environment, as we observed a lower mean number of somatic mutations in the SBS88 positive CRCs than in the SBS88 negative CRCs. Utilizing structural variants or CNA events resulting from colibactin-induced DNA damage in the mutational signature algorithm may help to refine the SBS88 mutational signature. In this study, 819 CRCs or 13.4% of the total 6,111 CRCs available for analysis could not be included in mutational signature calculation because of low (<5) somatic SNV count.

We observed a strong enrichment of SBS88 positive CRC in the distal colon and rectum. A previous study also found tumors exhibiting the colibactin signature more prevalent in distal tumors^18^. Differences in the prevalence of *pks^+^ E. coli* across the colon and rectum have been inconsistently reported^13, 14, 43^. Physiological differences across the colon and rectum are known to affect the composition and behavior of microbial populations^44^. The reported observations that *pks^+^ E. coli* occurs throughout the colon and rectum but SBS88 positive CRC is more prevalent in the distal colon and rectum suggests that colibactin has increased oncogenic potential in the mucosa of the distal colon and rectum. Furthermore, CRC in the distal colon is predominantly associated with development via the chromosomal instability pathway^45, 46^. In this study, SBS88 positive CRC was associated with specific chromosomal instability events, including copy number losses on 14q, and with recurrent somatic mutations in *APC, SMAD4* and *TP53* genes that are known driver genes of tumorigenesis via the chromosomal instability pathway. Therefore, the distal colon and rectum may provide more favorable conditions for colibactin-induced DNA damage that targets the necessary driver genes and CNAs to drive CRC development via the chromosomal instability pathway.

The observed heterogeneity within the SBS88 positive CRCs is an interesting finding. Three distinct subgroups of SBS88 positive CRC were identified, each characterized by an enrichment of different genomic features and location in the colon and rectum (**Supplementary Figure 11**). Clusters 1 and 2 were both predominantly found in the distal colon and rectum but differed by the presence of the *APC*:c.835-8A>G recurrent mutation (Cluster 1) and presence of *TP53* mutations and greater number of CNA events (cluster 2) while Cluster 3 was associated with the proximal colon, *KRAS* mutations, and an absence of recurrent hotspot mutations and CNA events. The timing, repetition, duration and strain of *pks^+^ E. coli* may drive this genomic heterogeneity. Shorter exposure duration and less genotoxic strains are associated more with structural variation such as interstrand cross-links and CNAs compared with SNVs and indels^16^, while the presence of the SBS88 signature in normal colonic mucosa^18^, in conjunction with the presence of the *APC*:c.835-8A>G recurrent somatic mutation in adenoma tissue^38^ and our finding that *APC*:c.835-8A>G is a likely early driver event, suggests early life exposure to colibactin may be important for CRC predisposition via *APC*:c.835-8A>G driven mechanism. Thus, the distinct genomic clusters may represent exposure-dependent pathways to tumorigenesis. Furthermore, the role of potential modifiers, including Western diet^13^, may contribute to the genomic heterogeneity and any exposure-dependent relationship with colibactin-induced SBS88 positive CRC.

The survival analysis indicated a better prognosis associated with SBS88-positive CRCs. The reason for this is unclear. CRC-specific survival has been linked with the immune response where immune infiltration is strongly associated with better prognosis^47^. *Pks^+^ E. coli* demonstrates complex interactions with the immune system^43^, suggesting *pks^+^ E. coli* infection impacts survival via its effect on the host’s immune response. We did not observe differential survival within the clusters, although increased copy number load is typically associated with poorer outcomes^48^.

The strengths of this study are the large sample size of targeted sequenced CRCs with associated clinico-pathological and survival data for CRC-specific death enabling adequately-powered analyses. The targeted capture was designed to capture genes important in CRC development (versus a pan-cancer designed panel) ensuring that identified targets or gene associations will be broadly relevant to future CRC diagnostics and/or treatment. This study has some limitations. It was performed on a targeted sequencing platform, which limits the feasibility of some genomic analyses. Due to the sequencing technology, the copy number analysis was not performed on the full dataset, thus reducing the available samples for this component of the analysis, which may have limited our ability to identify additional significantly associated CNA regions. In addition, panel-sequenced data does not allow base-level resolution of breakpoints which would enable us to confirm the surrounding genomic context of copy number related breakpoints to add confidence in a colibactin related damage profile. The ID18 signature could not be determined with accuracy in this study given the low number of indels; future studies utilizing whole-exome or whole-genome sequencing may enable further investigation of colibactin induced DNA damage and clinico-pathological and genomic features in CRC.

## Conclusions

The detection of tumorigenesis caused by colibactin-induced DNA damage from *pks^+^ E. coli* represents our ability, for the first time, to assign a non-hereditary etiology to any given CRC. This has important implications for the patient where assigning a cause for their cancer can relieve the anxiety of not otherwise knowing the cause after hereditary CRC genes have been excluded. The identification of this novel subtype of CRC will impact future opportunities for CRC prevention including via the detection of the SBS88 signature and/or the *APC:c.835- 8A>G* mutation as a biomarker of a current or prior *pks^+^ E. coli* infection and may additionally represent a biomarker of the malignant potential of adenoma or colonic mucosa, all of which may modify patient surveillance and management. Opportunities for prevention at the population level would conceivably include approaches that target the detection of *pks^+^ E. coli* in saliva or stool. Potential treatments that inhibit the genotoxic effects of colibactin are gaining momentum^49, 50^, underscoring the importance of detecting the SBS88 signature and/or the *APC:c.835-8A>G* mutation biomarker early. SBS88-positive CRCs were significantly associated with unique genomic alterations including the recurrent somatic mutation *APC:c.835-8A>G,* which is likely to be an early driver event. Extending this concept, it seems likely that there is a subset of genomic contexts throughout the genome that are both vulnerable to colibactin-induced DNA damage and that when mutated, drive CRC initiation and progression via the chromosomal instability pathway in the distal colon and rectum. Several knowledge gaps exist regarding the mechanisms driving genomic heterogeneity of SBS88 positive CRC, the timing of colibactin exposure, and potential modifiers that may increase oncogenic potential remain to be resolved. The findings from this study provide an important clinicopathological and genomic characterization of this novel subtype of CRC arising from a specific and likely modifiable gut bacteria and provide further elucidation of the mechanism underlying the colibactin-induced tumorigenesis and molecular phenotype associated with this CRC subgroup.

## Data Availability

The panel-sequenced data used in this study are available at the database of Genotypes and Phenotypes (dbGaP). The Ontario Institute of Cancer Research (OICR) data is available under accession code phs002050.v1.p1. The Center for Inherited Disease Research (CIDR) data is available under accession code phs001905.v1.p1. Mutational signature definitions were downloaded from the COSMIC website at https://cancer.sanger.ac.uk/signatures/downloads/.

https://www.ncbi.nlm.nih.gov/projects/gap/cgi-bin/study.cgi?study_id=phs002050.v1.p1

https://www.ncbi.nlm.nih.gov/projects/gap/cgi-bin/study.cgi?study_id=phs001905.v1.p1

https://cancer.sanger.ac.uk/signatures/downloads/

## Author Contributions

BJP, DDB, FAM, HB, IMW, JJ, MAJ, PG and UP developed the initial concept and design. AG, ATC, DDB, FAM, GGG, HB, IMW, JAN, JCC, JCF, JDP, JEM, MAJ, MH, MJG, MOS, MS, PTC, RES, RKP, SBG, SG, VM and WYH contributed to sample recruitment. AJF, ATC, CQ, DAD, DDB, JRH, KFD, MC, MJG, MOS, PTC, QMT, RS, RW, SBG, SHZ, SIB, SO, TAH, TJH, TU, UP, VM, WS, WYH and YL contributed to sample preparation and QC. AET, AIP, BJP, CYU, DAD, DDB, FAM, HB, IMW, JCF, JJ, KM, KT, MAJ, MG, PG, PTC, SO, TAH, TU, UP and YC helped interpret the findings. AIP, BJP, DDB, MAJ, PG and UP supervised the project. All authors read and approved the final manuscript.

## Data Availability Statement

The original panel-sequenced data used in this study are available at the database of Genotypes and Phenotypes (dbGaP). The Ontario Institute of Cancer Research (OICR) data is available under accession code <u>phs002050.v1.p1</u>. The Center for Inherited Disease Research (CIDR) data is available under accession code <u>phs001905.v1.p1</u>. Mutational signature definitions were downloaded from the COSMIC website at https://cancer.sanger.ac.uk/signatures/downloads/.

## ACKNOWLEDGEMENTS

CORSA: We kindly thank all individuals who agreed to participate in the CORSA study. Furthermore, we thank all cooperating physicians and students and the Biobank Graz of the Medical University of Graz.

CPS-II: The authors express sincere appreciation to all Cancer Prevention Study-II participants and to each member of the study and biospecimen management group. The authors would like to acknowledge the contribution to this study from central cancer registries supported through the Centers for Disease Control and Prevention’s National Program of Cancer Registries, and cancer registries supported by the National Cancer Institute’s Surveillance Epidemiology and End Results Program.

DACHS: We thank all participants and cooperating clinicians, and everyone who provided excellent technical assistance.

Harvard cohorts (HPFS, NHS): The study protocol was approved by the institutional review boards of the Brigham and Women’s Hospital and Harvard T.H. Chan School of Public Health, and those of participating registries as required. We would like to thank the participants and staff of the HPFS and NHS for their valuable contributions as well as the following state cancer registries for their help: AL, AZ, AR, CA, CO, CT, DE, FL, GA, ID, IL, IN, IA, KY, LA, ME, MD, MA, MI, NE, NH, NJ, NY, NC, ND, OH, OK, OR, PA, RI, SC, TN, TX, VA, WA, WY. The authors assume full responsibility for analyses and interpretation of these data.

PLCO: PLCO is supported by the Intramural Research Program of the Division of Cancer Epidemiology and Genetics and by contracts from the Division of Cancer Prevention, National Cancer Institute, NIH, DHHS. The authors thank the study management team at the Division of Cancer Epidemiology and Genetics and the Division of Cancer Prevention, National Cancer Institute (NCI), NIH, DHHS, staff at the Information Management Services, Inc., staff at Westat, Inc., staff at the Frederick National Laboratory for Cancer Research, Leidos Biomedical Research, Inc., and staff at the NCI Frederick Central Repository American Type Culture Collection (ATCC). Most importantly, we thank the study participants for their contributions that made this study possible. Cancer incidence data have been provided by the District of Columbia Cancer Registry, Georgia Cancer Registry, Hawaii Cancer Registry, Minnesota Cancer Surveillance System, Missouri Cancer Registry, Nevada Central Cancer Registry, Pennsylvania Cancer Registry, Texas Cancer Registry, Virginia Cancer Registry, and Wisconsin Cancer Reporting System. All are supported in part by funds from the Center for Disease Control and Prevention, National Program for Central Registries, local states or by the National Cancer Institute, Surveillance, Epidemiology, and End Results program. The results reported here and the conclusions derived are the sole responsibility of the authors.

SFCCR: The authors would like to thank the study participants and staff of the Seattle Colon Cancer Family Registry and the Hormones and Colon Cancer study (CORE Studies).

WHI: The authors thank the WHI investigators and staff for their dedication, and the study participants for making the program possible. A full listing of WHI investigators can be found at: http://www.whi.org/researchers/Documents%20%20Write%20a%20Paper/WHI%20Investigator%20Short%20List.pdf

