## Supplementary Methods for "Genotoxic colibactin mutational signature in colorectal cancer is associated with clinicopathological features, specific genomic alterations and better survival"

#### Study Participants

The study population included men and women diagnosed with incident invasive primary colon or rectal cancer (CRC) that were enrolled in one of the following participating studies (**Supplementary Table 1**):

***Colorectal Cancer Family Registry (CCFR)***

The CCFR consists of six centres dedicated to the establishment of a comprehensive collaborative infrastructure for interdisciplinary studies in the genetic epidemiology of CRC (1). The CCFR includes data from approximately 42,500 total subjects from 15,000 families (10,500 probands, and 26,770 unaffected and affected relatives and 4,276 unrelated controls and 923 spouse controls). Cases and controls, with ages ranging from 20 to 74 years, were recruited at the six participating centres beginning in 1998. Between 1999 and 2002, female cases and controls 50-74 years enrolled into the Seattle CCFR (SCCFR) were subsequently enrolled in a complementary study of post-menopausal hormone use and CRC risk (PMH). CCFR and PMH implemented a standardized questionnaire that is administered to all participants, and includes established and suspected risk factors for colorectal cancer, which includes questions on medical history and medication use, reproductive history (for female participants), family history, physical activity, demographics, alcohol and tobacco use, and dietary factors. The studies in this analysis selected tumour samples from population-based cases in the following population-based centres: Seattle (including a subset of PMH), Ontario (OFCCR), and Australia (AFCCR).

***Colorectal Cancer Study of Austria (CORSA)***

In the ongoing CORSA study, more than 16,000 Caucasian participants have been recruited within the province-wide screening project “Burgenland Prevention Trial of Colorectal Disease with

Immunological Testing” (B-PREDICT) since 2003 (2). All inhabitants of the Austrian province Burgenland aged between 40 and 80 years are annually invited to participate in faecal immunochemical testing and hemoccult positive screening participants are invited for colonoscopy. CORSA participants have been recruited in the four KRAGES hospitals in Burgenland, Austria, and additionally, at the Medical University of Vienna (Department of Surgery), the Viennese hospitals “Rudolfstiftung” and the “Sozialmedizinisches Zentrum Süd”, and at the Medical University of Graz (Department of Internal Medicine) (3).

***American Cancer Society Cancer Prevention Study II (CPSII)***

The CPS-II Nutrition cohort is a prospective study of cancer incidence and mortality in the United States, established in 1992 (4,5). At enrollment, participants completed a mailed self-administered questionnaire including information on demographic, medical, diet, and lifestyle factors. Follow-up questionnaires to update exposure information and to ascertain newly diagnosed cancers were sent biennially starting in 1997. Reported cancers were verified through medical records, state cancer registry linkage, or death certificates.

***Cancer Risk Assessment Study (CRA)***

All individuals who had surgery at the Mayo Clinic Rochester MN, Methodist Hospital Rochester MN, or St. Mary’s Hospital Rochester MN, who consented to participate from 1995 to 1998 were included in this study’s collection (6). Every patient was offered a chance to participate, no exclusion criteria were applied. Subjects who consented were given a form to complete that contained questions about lifestyle, medical history, and family history. Materials collected from subjects included peripheral blood, resected tumour from the centre and the rim of the neoplasm, as well as normal colon both adjacent to the tumour and at the surgical margin. One representative piece from each was flash frozen and stored at -70C. The remaining piece was fixed in formalin and embedded in paraffin. Additionally, if lymph nodes or metastatic tumours were present, material was collected for these as well.

*Colorectal Cancer Genetics & Genomics (CRCGEN, CRCGEN_2)*

This Spanish study combines data from three case-control studies. The first one, performed in University Hospital of Bellvitge, L'Hospitalet, Barcelona, recruited incident pathology-confirmed CRC cases during the period 1996-1998. The second study was performed in the same hospital during the period 2007-2015 and the third study was conducted in Hospital of Leon, Leon, during 2008-2013. This study included both colorectal cancer cases and adenoma cases. Adenoma or serrated polyps were detected at screening colonoscopy. Patients with high-risk lesions were selected, defined as ≥ 5 adenomas/serrated polyps, or ≥ 1 adenoma/serrated polyp ≥ 20mm.

*Darmkrebs: Chancen der Verhütung durch Screening (DACHS)*

This German study was initiated as a large population-based case-control study in 2003 in the Rhine-Neckar-Odenwald region (southwest region of Germany) to assess the potential of endoscopic screening for reduction of CRC risk and to investigate etiologic determinants of disease, particularly lifestyle/environmental factors and genetic factors (7). Cases with a first diagnosis of invasive CRC (International Classification of Diseases 10 codes C18-C20) who were at least 30 years of age (no upper age limit), German speaking, a resident in the study region, and mentally and physically able to participate in a one-hour interview, were recruited by their treating physicians either in the hospital a few days after surgery, or by mail after discharge from the hospital. Cases were confirmed based on histologic reports and hospital discharge letters following diagnosis of CRC. All hospitals treating CRC patients in the study region participated. Based on estimates from population-based cancer registries, more than 50% of all potentially eligible patients with incident CRC in the study region were included. During an in-person interview, data were collected on demographics, medical history, family history of CRC, and various life-style factors, as were blood and mouthwash samples. Formalin-fixed, paraffin-embedded, surgical specimens of CRC patients were collected from cooperating pathology institutes and transferred to the tissue bank of the National Center for Tumor Diseases in Heidelberg.

**European Prospective Investigation into Cancer and Nutrition, Norfolk site (EPIC)** (8)

The EPIC study is a multi-center, prospective cohort designed to investigate the associations between diet, cancer and other chronic diseases across 10 European countries: Denmark, France, Germany, Greece, Italy, the Netherlands, Norway, Spain, Sweden and the United Kingdom (UK). Participants were recruited between 1992 and 1998 and included 521,330 men and women aged 35–70 years. Details of this study have been previously described and are available online (https://www.epic-norfolk.org.uk/). For this study, FFPE tumors from the EPIC Norfolk site were selected for inclusion. All colorectal cancers were diagnosed and treated at the Norfolk and Norwich Hospital, Norwich, UK.

***Hispanic Colorectal Cancer Study (HCCS)***

HCCS is a population-based study of individuals self-identified as Hispanic with a diagnosis of colorectal cancer. Cases are identified from the California Cancer Registry or directly from local hospitals in the Los Angeles region [LAC + USC County Hospital and University of Southern California (USC) Norris Comprehensive Cancer Center]. All men and women over 21 years of age with a first-time diagnosis of CRC (ICD-O-3 codes: C18–C21) after January 1, 2008, were eligible for participation. Risk factor and dietary questionnaires, pathology reports, and saliva samples (for genotyping) were collected using methodologies developed in the Colon Cancer Family Registry and the Multiethnic Cohort (MEC). Participants recruited into the HCCS were born in Mexico, the US, Central/South America, Cuba, the Caribbean Islands, or Europe. The present study includes a number of cases with FFPE colorectal tumour tissue available.

***Health Professional's Follow-up study (HPFS)***

The HPFS cohort comprises over 51,000 men aged 40-75 years at enrolment and followed since the study started in 1986 (9,10). Participants provided information on health-related exposures, including current and past smoking history, weight, height, diet, supplement use, alcohol intake, physical activity, aspirin use, endoscopy procedures, and family history of colorectal cancer every two years (or four years for diet) through questionnaires. Colorectal cancer and other outcomes were reported by participants or next-of-kin and were followed up through review of the medical and pathology record by physicians. Lethal unreported colorectal cancer cases were identified (and confirmed) through next-of-kin, use of the National Death Index and medical record review. Overall, more than 97% of self-reported colorectal cancers were confirmed by medical record review. Information was abstracted on histology, primary tumour location, TNM staging, tumour size and multiplicity, and the number of positive and negative lymph nodes. In 1993–1995, over 18,000 participants mailed blood samples by overnight courier for buffy coat extraction and stored in liquid nitrogen. In 2001–2004, nearly 14,000 participants who had not provided a blood sample previously mailed in a swish-and-spit sample of buccal cells. FFPE tissue blocks were collected from hospitals where participants with colorectal carcinoma had undergone tumour resection or endoscopic biopsy (for pre-operatively treated rectal cancer). The study pathologist reviewed hematoxylin-and-eosin-stained tissue sections and recorded histopathological features.

***Iowa Women's Health Study (IWHS)***

In the IWHS, a 16-page baseline questionnaire was completed and returned by 41,836 randomly selected women, with ages ranging from 55 to 69 years, who resided in Iowa and held a valid driver's license at baseline in 1986 (11,12). Comprehensive self-reported demographic, dietary, lifestyle, and medication data were collected during the baseline IWHS evaluation (1986). Incident colorectal cancer cases were identified through annual linkage with the Iowa Cancer Registry, which is a member of the National Cancer Institute's Surveillance, Epidemiology, and End Results (SEER) program. Colorectal cancer cases were identified using International Classification for Diseases in Oncology (ICD-O) codes. Beginning in 2006, archived, paraffin-embedded tissue specimens were requested from incident colorectal cancer cases diagnosed through December 31, 2002.

***Melbourne Collaborative Cohort Study (MCCS)***

The MCCS is a prospective cohort study of 41,513 healthy adult volunteers between the ages of 27 and 76 years (99% aged 40-69) recruited from the Melbourne (Australia) metropolitan area between 1990 and 1994 (13). By 31 December 2009, 1,046 participants had a first histopathological diagnosis of invasive adenocarcinoma of the colon or rectum identified by a record linkage to the Victorian

Cancer Registry following the baseline study visit. Beginning in 2004, archived FFPE tissue specimens were requested from incident colorectal cancer cases diagnosed through 1995 to 2009. All CRC cases eligible for this study were selected based on the availability of a tumour sample and having no pre-baseline history of CRC (as confirmed by the Victorian Cancer Registry).
Melbourne Collaborative Cohort Study (MCCS) cohort recruitment was funded by VicHealth and Cancer Council Victoria. The MCCS was further augmented by Australian National Health and Medical Research Council grants 209057, 396414 and 1074383 and by infrastructure provided by Cancer Council Victoria. Cases and their vital status were ascertained through the Victorian Cancer Registry and the Australian Institute of Health and Welfare, including the Australian Cancer Database.

***Nurses' Health Study (NHS)***

The NHS cohort began in 1976 when over 121,000 female registered nurses ages 30 to 55 years returned the initial questionnaire that ascertained a variety of important health-related exposures (14). Colorectal cancer and other outcomes were reported by participants or next-of-kin and followed up through review of the medical and pathology record by physicians. Overall, more than 97% of self-reported colorectal cancers were confirmed by medical-record review. Participants have been sent questionnaires biennially to update information on lifestyle factors and newly diagnosed disease. Data on histology and primary location were abstracted. FFPE tissue blocks were collected from hospitals where participants with colorectal carcinoma had undergone tumour resection or endoscopic biopsy (for pre-operatively treated rectal cancer).

***Nurses' Health study II (NHSII)***

The Nurses' Health Study II is an ongoing cohort of over 116,000 female registered nurses in the US, aged 25-42 years at baseline in 1989 (15). Demographic, lifestyle and health-related information were obtained from participants at baseline and updated every two years using self-administered questionnaires. Study participants who had not previously reported a diagnosis of cancer and had responded to the 1995 study questionnaire were invited to provide blood samples between 1996 and

1999. Blood samples were collected from over 29,000 participants, aged 32 to 54 years at the time of blood draw. Similarly, between 2004 and 2006, active study participants who had not previously provided a blood sample were invited to provide buccal samples. Swish-and-spit samples of buccal cells were received from nearly 30,000 participants. Participants with a prior history of any cancer (except non-melanoma skin cancer), ulcerative colitis, or familial polyposis syndromes were excluded. FFPE tissue blocks were collected from hospitals where participants with colorectal carcinoma had undergone tumour resection or endoscopic biopsy (for preoperatively treated rectal cancer).

***Prostate, Lung, Colorectal, and Ovarian Cancer Screening Trial (PLCO)***

PLCO is a large, randomized, two-arm trial that enrolled over 154,000 men and women between the age of 55 and 74 years at ten centres in the United States in order to determine the effectiveness of screening to reduce cancer mortality (16,17). Half of the participants were randomized into the screening arm and half into the control arm. Participants in the screening arm received sigmoidoscopy screening at baseline and 3 or 5 years after enrollment; participants in the control arm received usual care. Enrollment began in 1993 and concluded in 2001. Both arms were followed for cancer incidence and mortality for at least 13 years from baseline. Details of this study have been previously described and are available online (https://prevention.cancer.gov/major-programs/prostate-lung-colorectal-and-ovarian-cancer-screening-trial and https://cdas.cancer.gov/plco/). In 2006, FFPE pathology tissue samples were collected from PLCO participants who developed selected cancers, including colorectal cancer (18).

***Women's Health Initiative (WHI)***

The WHI study is a large, multi-centre study of postmenopausal women aged 50 to 79 years at recruitment from 40 US clinical centres between 1993 and 1998, including over 68,000 women who participated in four overlapping trials evaluating: menopausal hormone therapy (HT: two trials), dietary modification (DM) and calcium-vitamin D (CaD) supplementation (19). Participants in the CaD trial were recruited from those who were either in the HT or the DM trial. Details of the WHI study design have been described elsewhere and are available online (20). FFPE pathology tissue samples were collected from WHI participants who developed selected cancers, including colorectal cancer. Patients with sufficient material and consent were included in this study.

**Targeted Sequencing**

Tumor DNA was extracted from FFPE sections and matching normal DNA from the blood, buccal, saliva, or adjacent normal colonic FFPE tissues was isolated. Tumor tissue was macrodissected from slides guided by an H&E (haematoxylin and eosin) stained slide marked for the tumor regions. All tumors underwent a pathology review to confirm that the tumor was a primary colorectal carcinoma. DNA was extracted from FFPE tissue using the QIAamp DNA Mini or QIAamp DNA FFPE tissue kits and normal DNA from other tissues using standard DNA extraction methods. DNA concentrations were determined by Quant-iT PicoGreen dsDNA Assay or the Qubit dsDNA HS Assay kits.

The amplicon panel-sequenced tumors and matched normal samples were processed as described previously (21). Briefly, DNA extracted from FFPE tissues was subjected to repair by using the PreCR Repair Mix (New England BioLabs, Ipswich, MA). AmpliSeq target amplification was performed using 20ng of genomic DNA for each of the two AmpliSeq primer pools. Following removal of primers, PCR products from each pool were combined and subjected to end repair and A-tailing using the KAPA HyperPrep Kit (Roche). Adapter ligation was performed using the NEXTflex DNA barcodes Kit (PerkinElmer) and libraries were analyzed on High Sensitivity TapeStation and submitted for cluster generation. Barcoded DNA sequence libraries were pooled using 48 samples for tumors and 48 or 192 samples for normal DNA. Paired-end sequencing was performed on HiSeq 2500 using the Illumina Genome Analyzer operating procedure. Paired-end reads were aligned to the reference human genome (GRCh37/hg19) using Burrows-Wheeler Aligner (BWA-MEM version 0.7.9a). Local realignments and base quality recalibrations were performed on aligned data. Only reads aligned uniquely to the reference human GRCh37/hg19 genome assembly were used in downstream analysis.

The hybridization capture panel sequenced tumors and matched normal samples were processed at the Center for Inherited Disease Research. The custom capture probes were designed internally at Integrated DNA Technologies (IDT) with the following tiling requirements: Coding Exons – 1x tiling across all regions, for small exon (<120bp) use 2 overlapping probes/exon; Additional content – 1x tiling across all regions; gene fusions – 2x tiling across all regions; bacterial genes – 1x tiling. Coding regions were run through BLAST, any probe with a hit >30 was filtered from the design. The final 1.96Mb panel contained the coding exons of 298 genes, 1205 regions of additional content, 5 gene fusions, 9 regions in bacterial genes and the CIDR barcode panel. A low input library protocol developed at CIDR (22) was performed. Libraries were prepared from 50- 200ng of genomic DNA, sheared for 80s using the Covaris LE220plus instrument (Covaris). The Kapa Hyper prep kit was used to process the sheared DNA into amplified dual-indexed adapter ligated fragments including a unique molecular index (UMI). All processing was done in 96 well plate formats using robotics (Beckman FXp, Perkin Elmer Janus, Agilent Bravo, Beckman NX). 'With Bead' clean ups were used following shearing and adapter ligation using GE Healthcare Sera-Mag Magnetic SpeedBeads (Carboxylate-Modified) beads. 750ng of amplified library was used in a pooled enrichment reaction (8 samples per normal pool, 4 samples per tumor pool) following IDT protocols (4hr hybridization). Repeats of initial failures were performed with 1 sample per enrichment reaction. Post-capture amplification was performed using the Kapa HiFi PCR enzyme with custom primers. Libraries were sequenced on the NovaSeq 6000 platform using 47 bp paired end runs and sequencing chemistry S2 Reagent Kit. NovaSeq flowcell data was demultiplexed using Picard (picard-2.17.6) ExtractIlluminaBarcodes followed by IlluminaBasecallsToSam. Output from IlluminaBasecallsToSam are read group level BAM files encoding the UMI sequence and quality score in RX and QX tags respectively. BAM files were converted to FASTQ, aligned with BWA Mem (version 0.7.17) to hg19 reference genome and merged with original BAM to add UMI sequence. Mapped and tagged reads were grouped by UMI sequence with fgbio's (version 1.2.0) GroupReadsByUmi module (strategy parameter set to adjacency, edit distance between UMIs set to 1, minimum mapping quality set to 20). Grouped reads were collapsed into molecular consensus reads (1 read per group minimum, minimum phred-scaled error rate 30, minimum input base quality 20). Molecular consensus reads were aligned to hg19 reference with BWA Mem and realigned with GATK (version 3.7) InDelRealigner module.

**Bioinformatics Pipelines and Analysis**

Somatic variants were generated from the panel-sequenced tumors as described previously (21). Somatic SNVs were called using Strelka v1.0.1547 and MuTect v1.1.748, retaining only variants reported by both callers. Additional filters were applied based on strand bias, minor allele frequency in Exome Aggregation Consortium (ExAC), read-depth, alternative read-depth, and clustered read position (21). Amplicon artifact filtering was applied to remove cases where mutant allele frequency varied across read clusters. Indel calls were obtained using majority votes from VarScan2 v2.4.349 (23), VarDict (Feb 2017) (24), and Strelka v1.0.1547 (25). After initial filtering of indels based on coverage and mutant allele frequency, background signals of alternative reads in normal samples were identified. A background filter was constructed from read counts from tumors and normal samples to remove indel calls in a subset of samples where signals were not significantly higher than background.

MSI status was called using mSINGS (26). A baseline reference was established using control samples from peripheral blood and for each of the 169 microsatellite loci included in our panel design, we quantified and compared the number of differently sized repeats in tumor samples to the baseline for the same locus. A locus was considered unstable if the number of mutated alleles exceeded the baseline reference by three times the standard deviation. To define an MSI positive tumor, we evaluated the fraction of unstable microsatellite loci out of the total number of loci analyzed as well as a qualitative separation of samples (a cutoff fraction of 10% unstable loci). As an additional way to validate calls, we compared classification of MSI status results for participants that had both existing tumor marker data and determined tumor characteristics from the targeted sequencing data. The classifications from orthogonal approaches were highly concordant with 98.6% concordance for the 1534 individuals with information on MSI status from both sources.

When referring to gene alterations, the associated transcripts are NM_000038 (*APC*), NM_005359 (*SMAD4*), NM_000546 (*TP53*), NM_006218 (*PIK3CA*) and NM_017969 (*IWS1*).

*Mutational signatures*

Signature fitting (27) was performed using the simulated annealing method previously described by SignatureEstimation (28), using the pre-defined set of COSMIC version 3.2 single base substitution (SBS) signatures (29). This set was restricted to the 18 signatures that have previously been observed in the Pan-Cancer Analysis of Whole Genomes (PCAWG) cohort of 59 whole-genome sequenced CRC tumors (30), including the known base excision repair signatures SBS18 and SBS36 associated with defective *MUTYH* (31), SBS30 associated with defective *NTHL1* (32,33), SBS11 associated with red meat consumption (34) and the pks+E.coli signature SBS88. The full list of included signature definitions: SBS1, SBS10a, SBS10b, SBS10c, SBS10d, SBS11, SBS15, SBS17a, SBS17b, SBS18, SBS28, SBS30, SBS36, SBS37, SBS40, SBS44, SBS5, SBS88. The reconstruction error was calculated as the cosine distance between the observed mutational context counts, and the predicted mutational context counts as computed from the calculated mutational signatures, a value bounded by 0 and 1, 0 indicating maximal similarity (35).

*Copy Number Alterations*

Focal (gene-level) copy number alterations were generated using the GATK 4.1 copy number caller. Medium-sized (10Mb) CNAs were generated from focal level CNAs, by segmenting the genome into 10Mb regions. Segments containing at least three focal CNA calls with concordant predictions were considered to constitute a medium-sized copy number change. Similarly, large-scale (chromosome arm) CNAs were generated from focal level CNAs, by considering all focal copy number changes across each chromosome arm and the mode of the copy number calls across the chromosome arm was assigned to that chromosome arm. Fisher's exact test was used to assess statistical significance between cases and controls.

**TABLES**

**Supplementary Table 1:** Tumors included from each study in each analysis. Clinico-pathological analysis was performed on microsatellite stable (MSS/L) tumors with at least five mutations. The percentage of SBS88-positive tumors is relative to those included in the clinic-pathological analysis.

| **Study** | **Total Tumors Available** | **SBS88-positive tumors (%)** | **Clinico-Pathological Analysis** | **Survival Analysis** | **CNA Analysis** |
|---|---|---|---|---|---|
| ACCFR | 343 | 24 (8.5) | 282 | 273 | 343 |
| CORSA | 174 | 11 (8.6) | 128 | | |
| CPSII | 541 | 29 (8.9) | 326 | 311 | |
| CRA | 330 | 18 (7.2) | 249 | | 330 |
| CRCGEN | 796 | 42 (6.8) | 622 | 555 | 756 |
| DACHS | 278 | 8 (3.7) | 214 | 208 | |
| EPIC | 193 | 18 (12.6) | 143 | 51 | 193 |
| HCCS | 123 | 8 (9.2) | 87 | | 117 |
| HPFS | 300 | 13 (6.1) | 214 | 177 | 288 |
| IWHS | 393 | 32 (12.1) | 265 | 254 | 392 |
| MCCS | 475 | 39 (9.7) | 401 | 364 | 475 |
| NHS | 435 | 23 (8.6) | 269 | 252 | 422 |
| NHSII | 45 | 3 (10.3) | 29 | 29 | 42 |
| OCCFR | 685 | 63 (12.5) | 505 | 444 | |
| PLCO | 136 | 7 (7.5) | 94 | 87 | 136 |
| SCCFR | 529 | 32 (10.3) | 310 | 277 | |
| WHI | 335 | 22 (12.9) | 170 | | |
| **TOTAL** | **6,111** | **392 (9.1)** | **4,308** | **3,282** | **2,654** |

**Supplementary Table 2:** Comparison of genomic features across MSS/L CRC (n=4,308) stratified by SBS88 negative and SBS88 positive CRCs. P-values were either calculated with Fisher exact (2x2 categorical), or from a two-sided unpaired t-test (age of diagnosis). Statistically significant p-values (<0.05) are highlighted in bold.

| **Feature** | **SBS88 negative CRC (n=3,916)** | **SBS88 positive CRC (n=392)** | **p-value** | **Odds Ratio (95% CI)** |
|---|---|---|---|---|
| *Somatic CRC driver gene mutations* | | | | |
| *APC truncated first 1600AA* | | | | |
| Absent | 1,121 (88.7%) | 143 (11.3%) | | |
| Present | 2,795 (91.8%) | 249 (8.1%) | **$1x10^{-3}$** | **0.70 (0.56-0.87)** |
| *KRAS oncogenic known* | | | | |
| Absent | 2,224 (90.1%) | 244 (9.9%) | | |
| Present | 1,692 (92.0%) | 148 (8.0%) | **0.04** | 0.80 (0.64-0.99) |
| *NRAS oncogenic known* | | | | |
| Absent | 3753 (91.1%) | 365 (8.9%) | | |
| Present | 163 (85.8%) | 27 (14.2%) | **0.01** | **1.70 (1.12-2.60)** |
| *BRAF c.1799T>A (p.V600E)* | | | | |
| Absent | 3,697 (90.7%) | 381 (9.3%) | | |
| Present | 219 (95.2%) | 11 (4.8%) | **0.02** | 0.49 (0.26-0.90) |
| *PIK3CA nonsynonymous* | | | | |
| Absent | 3,281 (90.3%) | 353 (9.7%) | | |
| Present | 635 (94.2%) | 39 (5.8%) | **$1x10^{-3}$** | **0.57 (0.41-0.80)** |
| *PIK3R1 nonsilent* | | | | |
| Absent | 3,801 (90.9%) | 380 (9.1%) | | |
| Present | 115 (90.6%) | 12 (9.4%) | 0.89 | 1.04 (0.57-1.91) |
| *PTEN nonsilent* | | | | |
| Absent | 3,753 (90.7%) | 387 (9.3%) | | |
| Present | 163 (97.0%) | 5 (3.0%) | **$5x10^{-3}$** | **0.30 (0.12-0.73)** |
| *RNF43 truncating* | | | | |
| Absent | 3,832 (90.8%) | 387 (9.2%) | | |
| Present | 84 (94.4%) | 5 (5.6%) | 0.25 | 0.59 (0.24-1.46) |
| *TP53 nonsilent* | | | | |
| Absent | 1,412 (90.4%) | 150 (9.6%) | | |
| Present | 2,504 (91.2%) | 242 (8.8%) | 0.39 | 0.91 (0.73-1.13) |
| *Oncogenic Pathways* | | | | |
| *IGF2 PIK3 pathway* | | | | |
| Absent | 3,100 (90.2%) | 336 (9.8%) | | |
| Present | 816 (93.6%) | 56 (6.4%) | **$2x10^{-3}$** | **0.63 (0.47-0.85)** |
| *CTNNB1 hotspot* | | | | |
| Absent | 3,867 (90.9%) | 389 (9.1%) | 0.40 | |

| | | | | |
|---|---|---|---|---|
| Present | 49 (94.2%) | 3 (5.8%) | | 0.61 (0.19-1.96) |
| *TP53 pathway* | | | | |
| Absent | 1,264 (90.5%) | 133 (9.5%) | | |
| Present | 2,652 (91.1%) | 259 (8.9%) | 0.51 | 0.93 (0.75-1.16) |
| *RTK RAS pathway* | | | | |
| Absent | 1,690 (89.7%) | 195 (10.3%) | | |
| Present | 2,226 (91.9%) | 197 (8.1%) | **0.01** | **0.77 (0.62-0.94)** |
| *TGFβ pathway* | | | | |
| Absent | 2,902 (90.1%) | 319 (9.9%) | | |
| Present | 1,014 (93.3%) | 73 (6.7%) | **$2x10^{-3}$** | **0.65 (0.50-0.85)** |
| *WNT pathway* | | | | |
| Absent | 611 (88.2%) | 82 (11.8%) | | |
| Present | 3,305 (91.4%) | 310 (8.6%) | **$6x10^{-3}$** | **0.70 (0.54-0.90)** |

**Supplementary Table 3:** Stratified analysis of clinico-pathological features comparing SBS88-negative tumors with SBS88-positive tumors with and without the recurrent somatic variant *APC*:c.835-8A>G. Significant results (p-value<0.05) are in bold.

| **Feature** | **SBS88 negative (n=3916)** | **SBS88 positive without APCc.835-8A>G mutation (n=301)** | **Odds ratio (95%CI)** | **P-value** | **SBS88 positive with APCc.835-8A>G mutation (n=91)** | **Odds Ratio (95% CI)** | **P-value** |
|---|---|---|---|---|---|---|---|
| *Age at CRC diagnosis (mean±SD)* | 66.0 ± 11.7 | 64.8 ± 12.0 | n/a | 0.1 | 63.9 ± 11.6 | n/a | 0.1 |
| *Sex*<br>*Male*<br>*Female* | <br>1932 (49%)<br>1984 (51%) | <br>127 (42%)<br>174 (58%) | 1.3 (1.05-1.69) | **0.02** | <br>39 (43%)<br>52 (57%) | 1.3 (0.85-1.98) | 0.2 |
| *Tumor site*<br>*Proximal*<br>*Distal*<br>*Rectal* | <br>1400 (39%)<br>1176 (32%)<br>1057 (29%) | <br>84 (30%)<br>91 (33%)<br>103 (37%) | <br>Ref<br>1.3 (0.9-1.8)<br>1.6 (1.2-2.2) | **$6x10^{-3}$** | <br>8 (9%)<br>51 (58%)<br>29 (33%) | <br>Ref<br>7.6 (3.6-16.1)<br>4.8 (2.2-10.5) | **$8x10^{-9}$** |
| *Tumor stage*<br>*1*<br>*2*<br>*3*<br>*4* | <br>720 (24%)<br>890 (30%)<br>921 (31%)<br>449 (15%) | <br>54 (24%)<br>61 (27%)<br>85 (37%)<br>30 (13%) | <br>Ref<br>0.9 (0.6-1.3)<br>1.2 (0.9-1.8)<br>0.9 (0.6-1.4) | 0.27 | <br>20 (30%)<br>21 (31%)<br>17 (25%)<br>9 (13%) | <br>Ref<br>0.8 (0.5-1.6)<br>0.7 (0.3-1.3)<br>0.7 (0.3-1.6) | 0.64 |
| *Family history*<br>*No*<br>*Yes* | <br>2648 (83%)<br>550 (17%) | <br>200 (79%)<br>52 (21%) | <br>Ref<br>1.3 (0.9-1.7) | 0.17 | <br>60 (81%)<br>14 (19%) | <br>Ref<br>1.1 (0.6-2.0) | 0.70 |
| *# family relatives* | 0.2 ± 0.5 | 0.2 ± 0.5 | n/a | 0.07 | 0.2 ± 0.5 | n/a | 0.39 |
| *Ethnicity*<br>*White*<br>*Amer Ind.*<br>*Asian*<br>*Black/Afr*<br>*Other* | <br>3378 (97%)<br>2 (0.05%)<br>29 (0.8%)<br>33 (0.9%)<br>27 (0.8%) | <br>267 (97%)<br>0 (0%)<br>1 (0.3%)<br>4 (1%)<br>2 (0.7%) | <br>Ref<br>-<br>0.4 (0-3.2)<br>1.5 (0.5-4.4)<br>0.9 (0.2-4.0) | 0.82 | <br>83 (97%)<br>0 (0%)<br>1 (1%)<br>0 (0%)<br>2 (2%) | <br>Ref<br>-<br>1.4 (0.2-10.4)<br>-<br>3.0 (0.7-12.9) | 0.49 |
| *IBD*<br>*No*<br>*Yes* | <br>2331 (98%)<br>47 (2%) | <br>173 (98%)<br>4 (2%) | <br>Ref<br>1.1 (0.4-3.2) | 0.79 | <br>52 (100%)<br>0 (0%) | <br>Ref<br>0.9 (0.1-6.8) | 0.31 |

**Supplementary Table 4:** Counts of significant copy number alterations (CNAs) enriched in either SBS88-positive CRC or SBS88-negative CRC by three different CNA size categories (gene/focal level, 10Mb level or chromosome arm level) and by CNA type (loss or gain).

| CNA Type | Gain or Loss | No. Events Observed |
|---|---|---|
| Enriched in SBS88-positive CRC | | |
| Focal | Loss | 44 |
| Focal | Gain | 71 |
| 10Mb | Loss | 7 |
| 10Mb | Gain | 9 |
| Arm | Loss | 1 |
| Arm | Gain | 3 |
| Enriched in SBS88-negative CRC | | |
| Focal | Loss | 97 |
| Focal | Gain | 14 |
| 10Mb | Loss | 17 |
| 10Mb | Gain | 0 |
| Arm | Loss | 3 |
| Arm | Gain | 1 |

**Supplementary Table 5:** Medium-sized (10Mb) and the corresponding chromosome arm-level copy number alterations (CNAs) that were either significantly enriched or underrepresented in SBS88 positive CRCs compared with SBS88 negative CRCs. A total of 33 10Mb regions were significantly different, forming 17 contiguous regions on the genome. P-values were calculated with Fisher's exact test.

| **Arm-level CNAs** | **Medium-size (10Mb) CNAs** | **SBS88 negative (Control) (n=2,427)** | **SBS88 positive (Colibactin-induced CRC) (n=227)** | **p-value** | **Odds Ratio (95% CI)** |
|---|---|---|---|---|---|
| *Gene level (focal) CNAs* | | | | | |
| | **Median number of CNA gains (interquartile range)** | **191 (121-272)** | **201 (131.5-282)** | | n/a |
| | **Median number of CNA losses (interquartile range)** | **147 (64-244)** | **148 (78.5-244)** | | n/a |
| *CNAs enriched in SBS88 positive CRC* | | | | | |
| None | Loss chr1:30-40M<br>Absent<br>Present | <br>1904 (92.1%)<br>523 (89.2%) | <br>164 (7.9%)<br>63 (10.8%) | **0.04** | **1.4 (1.0-1.9)** |
| Gain 13q | Gain chr13:20-50M<br>Absent<br>Present | <br>728 (93.3%)<br>1699 (90.7%) | <br>52 (6.7%)<br>175 (9.3%) | **0.03** | **1.4 (1.0-2.0)** |
| | Gain chr13:70-80M<br>Absent<br>Present | <br>750 (93.6%)<br>1677 (90.5%) | <br>51 (6.4%)<br>176 (9.5%) | **0.01** | **1.5 (1.1-2.1)** |
| | Gain chr13:100-110M<br>Absent<br>Present | <br>751 (93.6%)<br>1676 (90.5%) | <br>51 (6.4%)<br>176 (9.5%) | **0.01** | **1.5 (1.1-2.1)** |
| Loss 14q | Loss chr14:20-40M<br>Absent<br>Present | <br>1674 (92.5%)<br>753 (89.1%) | <br>135 (7.5%)<br>92 (10.9%) | **$4x10^{-3}$** | **1.5 (1.1-2.0)** |
| | Loss chr14:50-70M<br>Absent<br>Present | <br>1679 (92.5%)<br>748 (89.2%) | <br>136 (7.5%)<br>91 (10.8%) | **$5x10^{-3}$** | **1.5 (1.1-2.0)** |
| | Loss chr14:90-110M<br>Absent<br>Present | <br>1696 (92.6%)<br>731 (88.9%) | <br>136 (7.4%)<br>91 (11.1%) | **$2x10^{-3}$** | **1.6 (1.2-2.0)** |
| Gain 16q | Gain chr16:50-80M<br>Absent<br>Present | <br>1942 (92.1%)<br>485 (89.0%) | <br>167 (7.9%)<br>60 (11.0%) | **0.03** | **1.4 (1.1-2.0)** |

| | | | | | |
|---|---|---|---|---|---|
| Gain 20p | Gain chr20:10-20M<br>Absent<br>Present | <br>1284 (92.6%)<br>1143 (90.2%) | <br>103 (7.4%)<br>124 (9.8%) | **0.03** | **1.4 (1.0-1.8**) |
| *CNAs underrepresented in SBS88 positive CRC* | | | | | |
| Loss 2p | Loss chr2:10-40M<br>Absent<br>Present | <br>2362 (91.3%)<br>65 (98.5%) | <br>226 (8.7%)<br>1 (1.5%) | **0.04** | **0.2 (0.02-1.0)** |
| | Loss chr2:70-80M<br>Absent<br>Present | <br>2383 (91.3%)<br>44 (100%) | <br>227 (8.7%)<br>0 (0%) | **0.03** | - |
| Loss 2q | Loss chr2:110-130M<br>Absent<br>Present | <br>2385 (91.3%)<br>42 (100%) | <br>227 (8.7%)<br>0 (0%) | **0.02** | - |
| | Loss chr2:150-170M<br>Absent<br>Present | <br>2381 (91.3%)<br>46 (100%) | <br>227 (8.7%)<br>0 (0%) | **0.03** | - |
| | Loss chr2:180-200M<br>Absent<br>Present | <br>2382 (91.3%)<br>45 (100%) | <br>227 (8.7%)<br>0 (0%) | **0.03** | - |
| | Loss chr2:210-240M<br>Absent<br>Present | <br>2371 (91.3%)<br>56 (100%) | <br>227 (8.7%)<br>0 (0%) | **0.01** | - |
| None | Loss chr12:0-10M<br>Absent<br>Present | <br>2287 (91.2%)<br>140 (95.9%) | <br>221 (8.8%)<br>6 (4.1%) | **0.05** | **0.4 (0.2-1.0)** |
| Loss 17q | Loss chr17:50-80M<br>Absent<br>Present | <br>2079 (90.9%)<br>348 (94.6%) | <br>207 (9.1%)<br>20 (5.4%) | **0.02** | **0.6 (0.4-0.9)** |

**Supplementary Table 6:** Summary of significant ($p<0.05$) CNA calls across 10Mb genomic regions and chromosomal arms. Grouped by CNA type (loss or gain) and chromosome, both contiguous regions of 10Mb regions of significance (medium-scale CNAs), and entire chromosome arms (large-scale CNAs) were found to have significant differences in prevalence in SBS88-positive and SBS88-negative tumors. Survival p-values based on adjustment for age, sex and cohort. Significant results (p-value $<0.05$) are in bold.

| **CNA type** | **Chromosome** | **Medium-scale CNAs (10Mb bins)** | **Large-scale CNAs (chromosome arm)** | **Large scale CNA survival p-value (odds ratio)** |
|---|---|---|---|---|
| Enriched in SBS88-positive CRC | | | | |
| Loss | 1 | 30-40Mb | None | |
| Loss | 14 | 20-40Mb<br>50-70Mb<br>90-110Mb | q | 0.10<br>(0.69-1.03) |
| Gain | 13 | 20-50Mb<br>70-80Mb<br>100-110Mb | q | 0.50<br>(0.88-1.31) |
| Gain | 14 | 50-70Mb<br>90-110Mb | None | |
| Gain | 16 | 50-80Mb | q | 0.06<br>(0.99-1.51) |
| Gain | 20 | 10-20Mb | p | 0.37<br>(0.91-1.30) |
| Enriched in SBS88-negative CRC | | | | |
| Loss | 2 | 10-40Mb<br>70-80Mb | p | Insufficient data |
| Loss | 2 | 110-130Mb<br>150-170Mb<br>180-200Mb<br>210-240Mb | q | 0.19<br>(0.82-2.71) |
| Loss | 12 | 0-10Mb | None | |
| Loss | 17 | 50-80Mb | q | **0.04<br>(1.01-1.61)** |
| Gain | 15 | None | q | **0.002<br>(1.33-3.44)** |

**Supplementary Table 7**: Stratification of 227 SBS88-positive tumors based on enriched genomic features (copy number alterations and *APC*:c.835-8 A>G).

| | **Cluster 1** | **Cluster 2** | **Cluster 3** |
|---|---|---|---|
| Cluster name | APC hotspot/20p gain cluster | TP53/CNA dominant cluster | WNT/CNA rare cluster |
| Proportion of SBS88 positive CRCs per cluster | 41.4%<br>n=94 | 33.0%<br>n=75 | 25.6%<br>n=58 |
| Genomic Features | ● *APC*:c.835-8 A>G hotspot<br>● 20p gain | ● *TP53* mutations<br>● 13q gain<br>● 14q loss<br>● 16q gain | ● *KRAS* mutations<br>● *TGFβ* pathway<br>● WNT pathway<br>● Absence of SBS88 associated CNAs<br>● Absence of *APC* recurrent hotspots |
| Anatomical site | Distal/rectum | Distal/rectum | Proximal |
| Mean (±SD) SBS88 proportion* | 21.4% (11.3%) | 19.8% (8.3%) | 17.4% (5.9%) |
| Mean (±SD) TMB+ | 5.4 (2.2) | 5.3 (2.0) | 6.1 (2.2) |
| Female frequency | 59.6% | 60% | 62.1% |
| Mean (±SD) Age at CRC diagnosis+ | 65.7 (10.6) | 64.4 (12.5) | 65.2 (11) |

*means of SBS88 proportion were significantly different between the three clusters (p=0.035)
+means of TMB and age at CRC diagnosis were not significantly different between the three clusters (p=0.058 and p=0.76, respectively)

**Supplementary Table 8:** Comparing numeric features across genomically determined clusters. Significant results (p-value<0.05) are in bold.

| | **Cluster 1** | **Cluster 2** | **Cluster 3** | |
|---|---|---|---|---|
| **Covariate** | **APC:c.835 C>A 20p GAIN** | **TP53 16q GAIN** | ***KRAS*** ***TGFβ*** **WNT CNAs rare** | **P-value** |
| Signature reconstruction error | 0.377 (0.119) | 0.380 (0.107) | 0.344 (0.123) | 0.158 |
| SBS88 proportion | 0.214 (0.113) | 0.198 (0.083) | 0.174 (0.059) | **0.035** |
| Age at CRC diagnosis | 65.681 (10.605) | 64.373 (12.475) | 65.175 (10.958) | 0.761 |
| No. of somatic INDELs | 0.968 (0.978) | 0.920 (0.983) | 1.293 (1.043) | 0.072 |
| No. of somatic SNVs | 9.628 (4.258) | 9.453 (3.761) | 10.707 (4.272) | 0.176 |
| TMB | 5.397 (2.146) | 5.284 (1.980) | 6.113 (2.240) | 0.058 |

### FIGURES

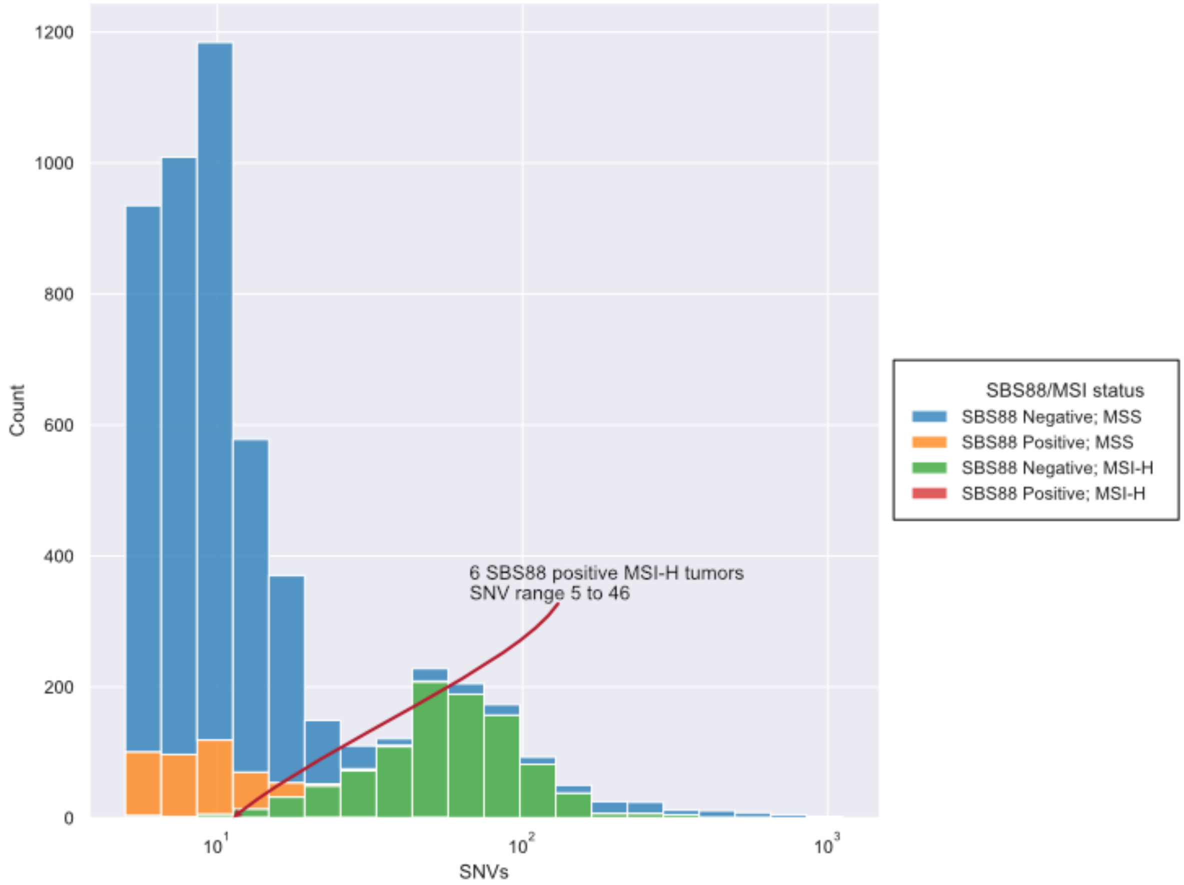


**Supplementary Figure 1**: Distribution of SBS88 positive and negative tumors by their MSS/L and MSI-H status across the SNV count for 4,308 CRCs. Tumors with a minimum of five somatic mutations are shown.

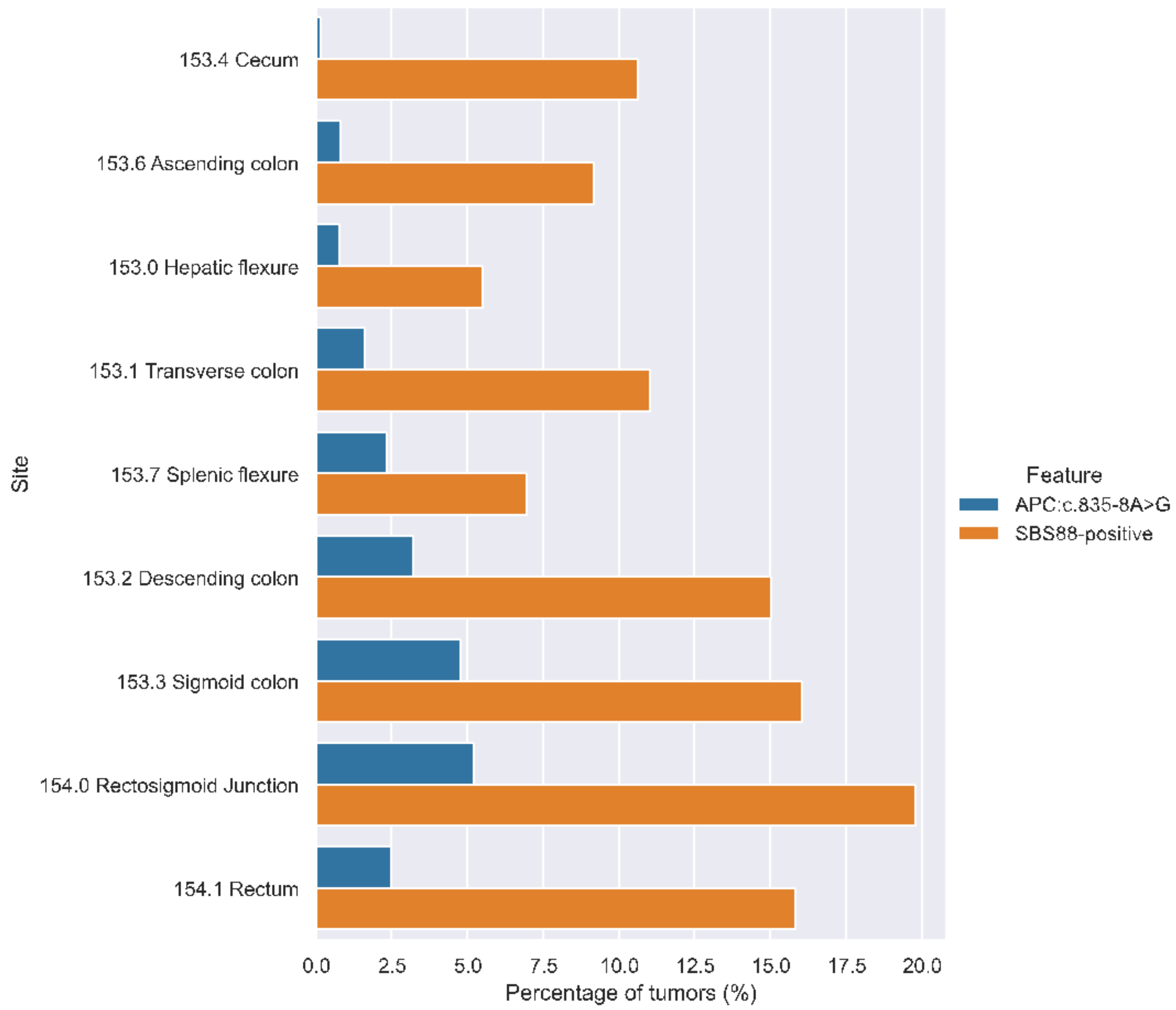


**Supplementary Figure 2:** Prevalence of SBS88-positive CRC, and the strongest associated somatic hotspot mutation, *APC*:c.835-8A>G, stratified by ICD tumor site for 4,308 CRCs.

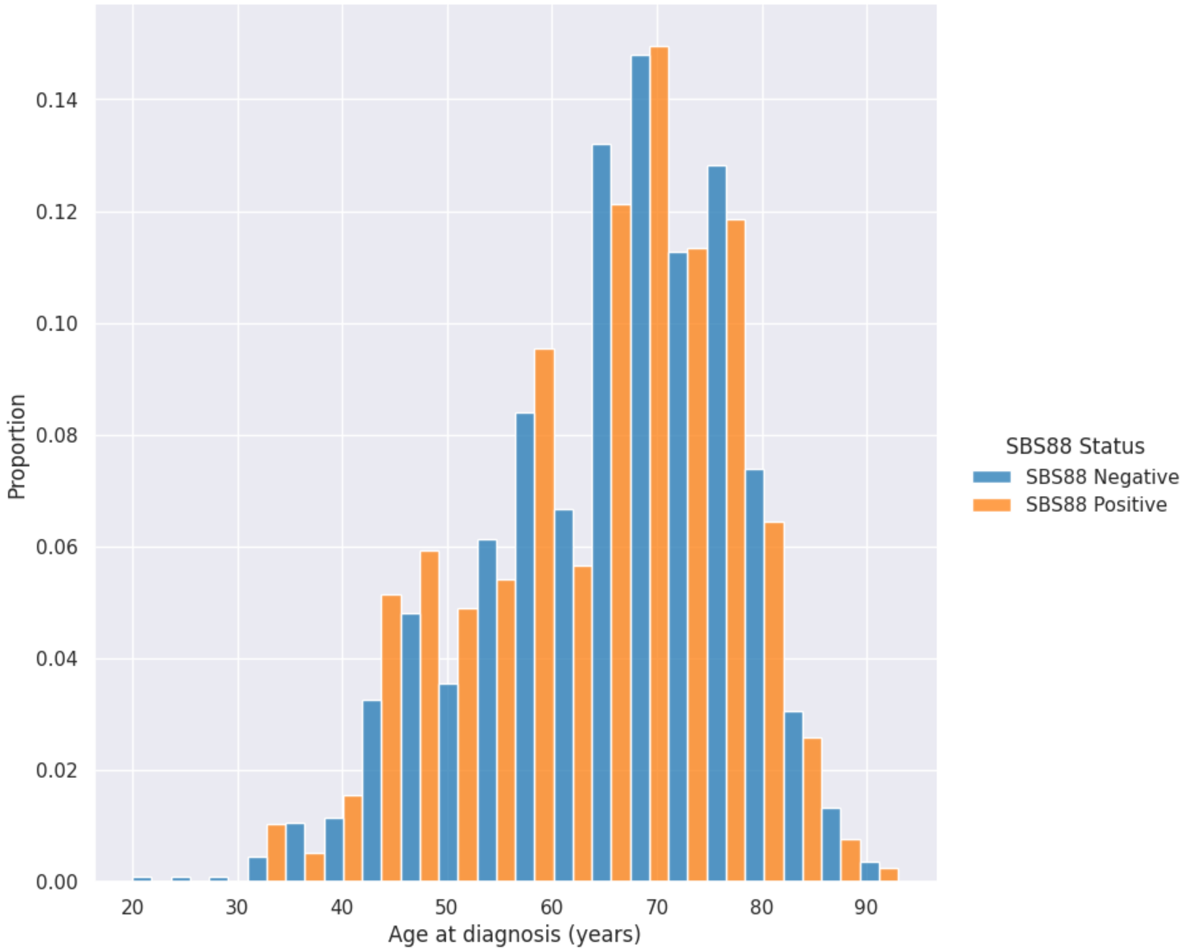


**Supplementary Figure 3:** Number of SBS88-positive and -negative CRCs relative to age at CRC diagnosis. SBS88-positive tumors showed a younger age of diagnosis (p=0.03; t-test).

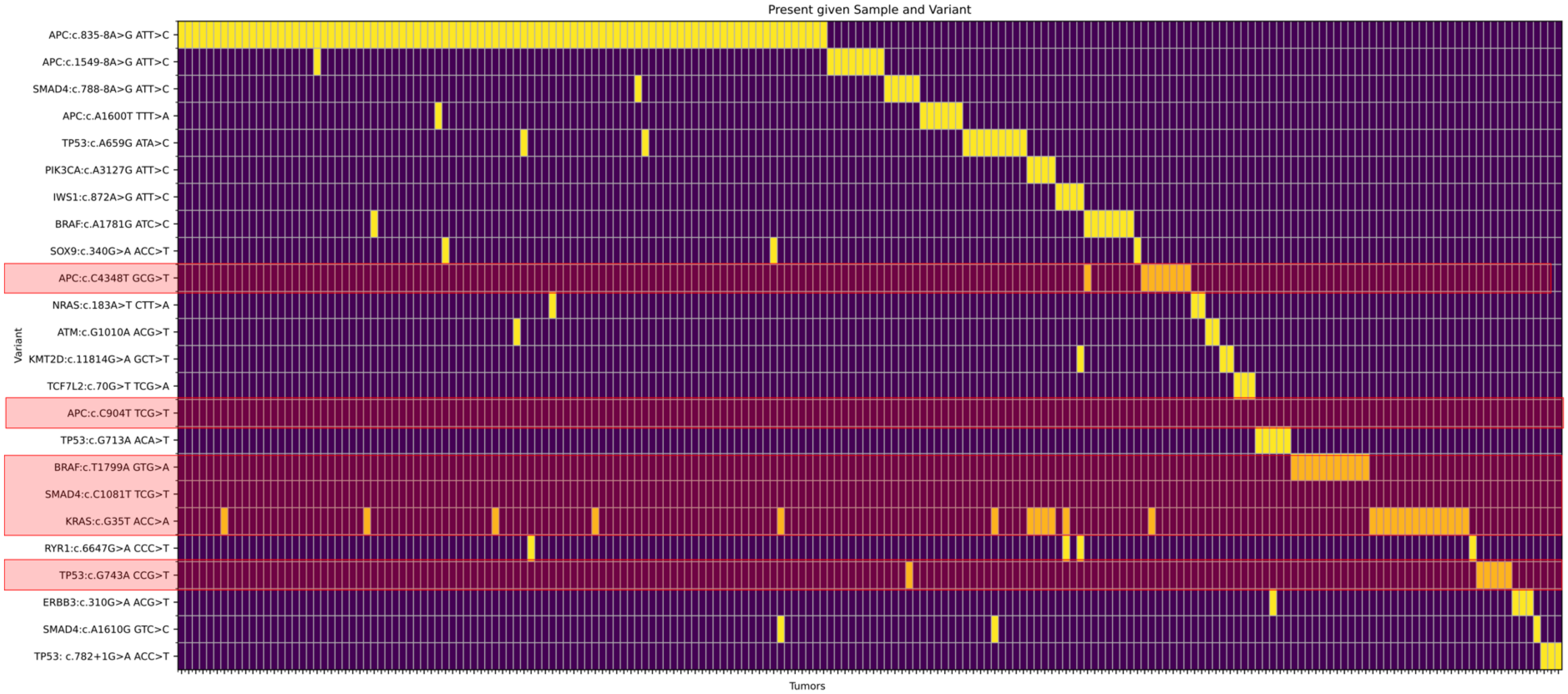


**Supplementary Figure 4:** Recurrent somatic mutations significantly associated with SBS88-positive CRC (n=24) where the majority (n=18) of these associated recurrent mutations were positively correlated (enriched) with SBS88-positive CRC while six recurrent mutations were significantly negatively correlated with SBS88-positive CRC (highlighted in red).

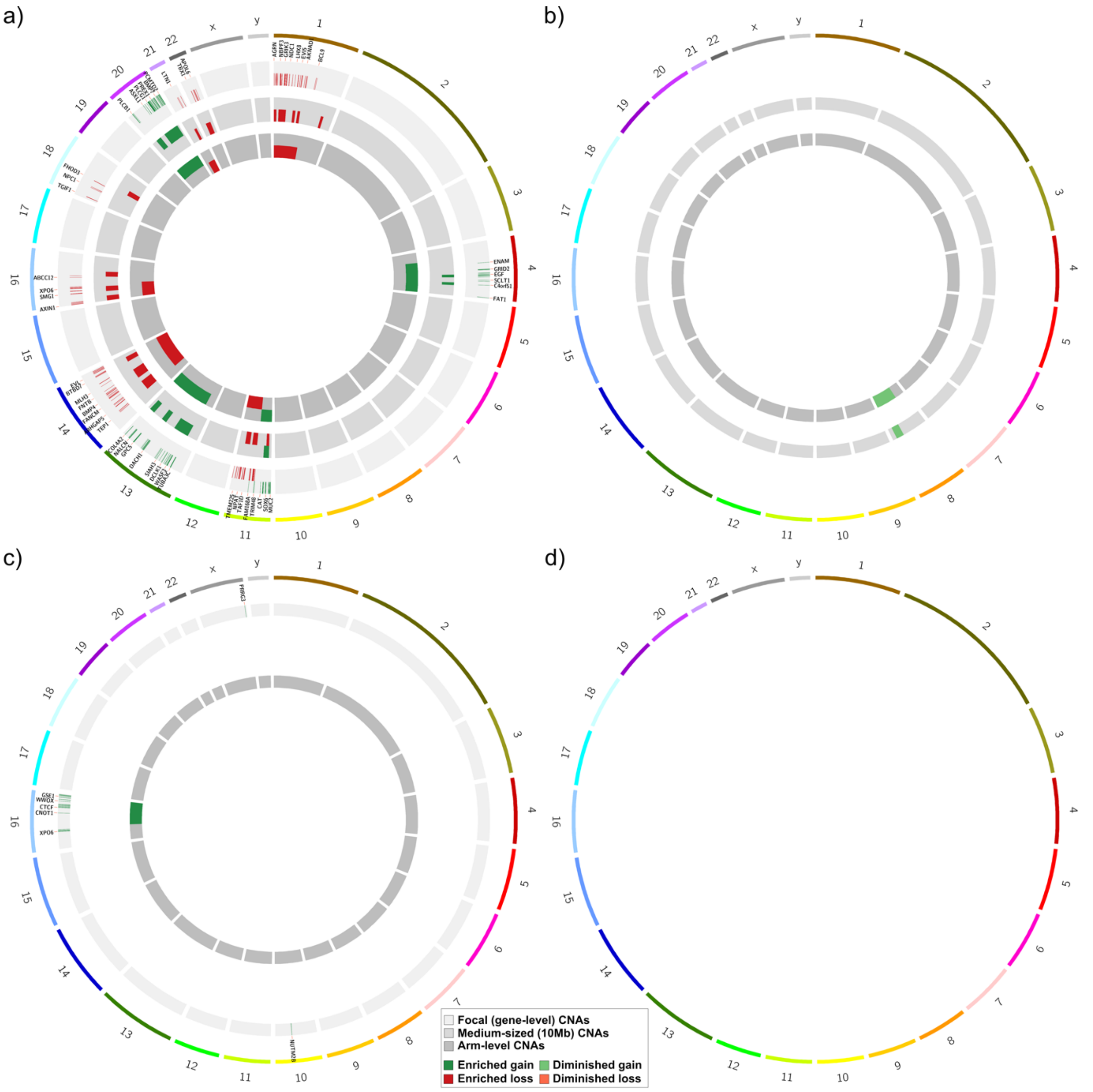


**Supplementary Figure 5:** Circos plots showing gene, medium and chromosome level CNA gains and losses that are significantly enriched in either (**a**) SBS88-positive tumors with the *APC*:c.835-8A>G mutation compared with SBS88-negative tumors (n=289 gene, n=39 medium and n=10 chromosome significant CNA events); or (**b**) SBS88-negative tumors compared with SBS88-positive tumors with *APC*:c.835-8A>G (n=3 medium and n=1 chromosome); or (**c**) SBS88-positive tumors without *APC*:c.835-8A>G compared to SBS88-negative tumors (n=29 gene and n=1 chromosome); or (**d**) SBS88-negative tumors compared with SBS88-positive tumors without *APC*:c.835-8A>G (no significant gene, medium and/or chromosome arm CNAs identified).

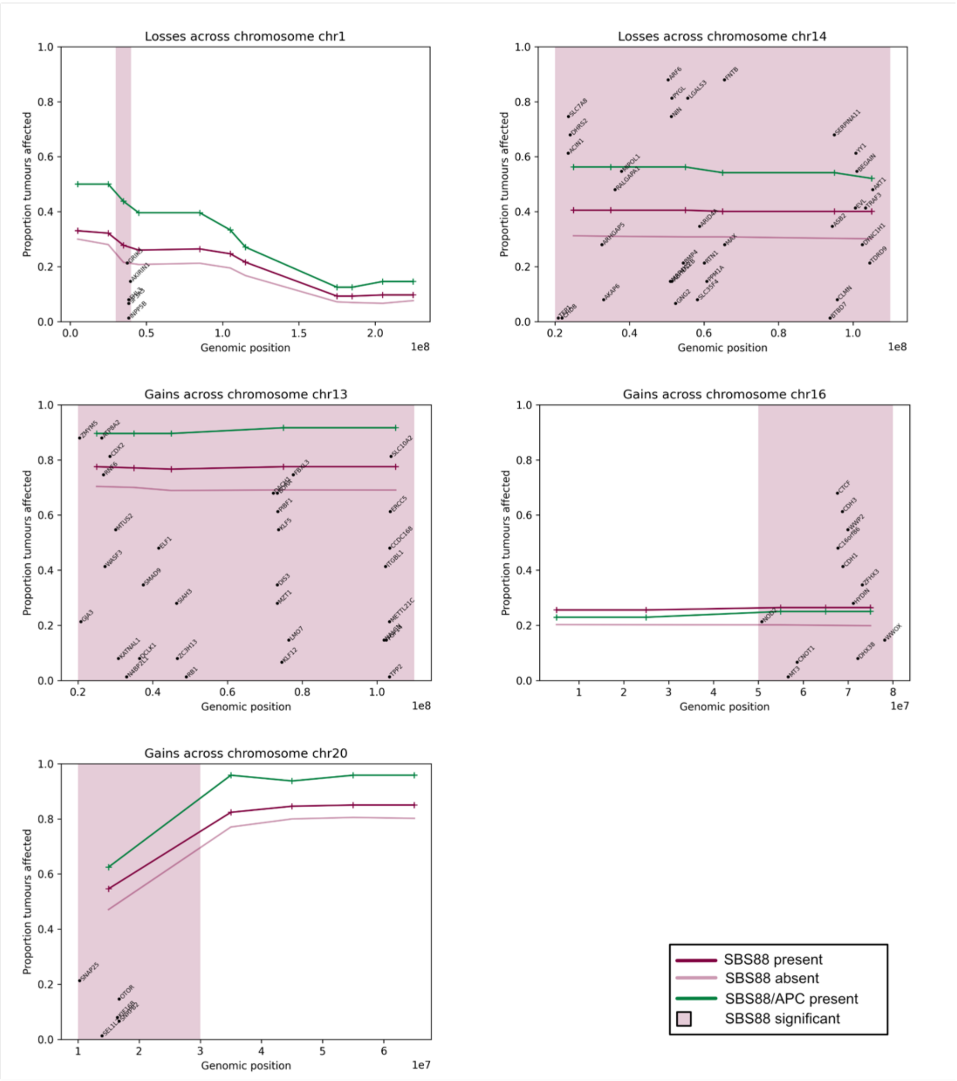


**Supplementary Figure 6:** Based on observed medium-sized (10Mb) CNAs, nine contiguous segments across five chromosome regions exhibited significant CNA differences between SBS88-negative and SBS88-positive CRCs (shaded light red). Tumors additionally showing the *APC*:c.835-8A>G hotspot mutation tended to be more likely to be affected, but less significant due to lower numbers. Genes in affected areas and covered by the sequencing panel are shown.

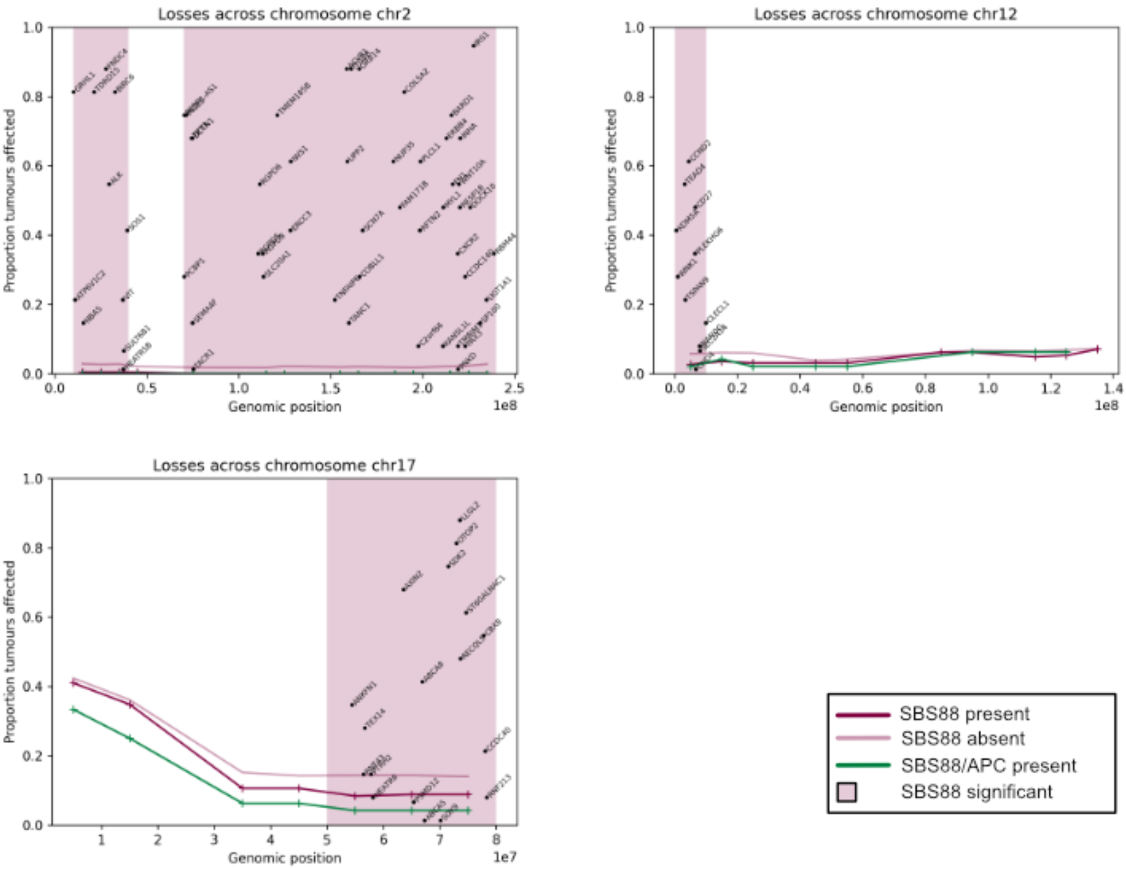


**Supplementary Figure 7:** Four contiguous regions showed significant CNA losses (red shading) in SBS88 negative tumors, compared to SBS88 positive tumors, based on the observed medium-sized (10Mb) CNA changes.

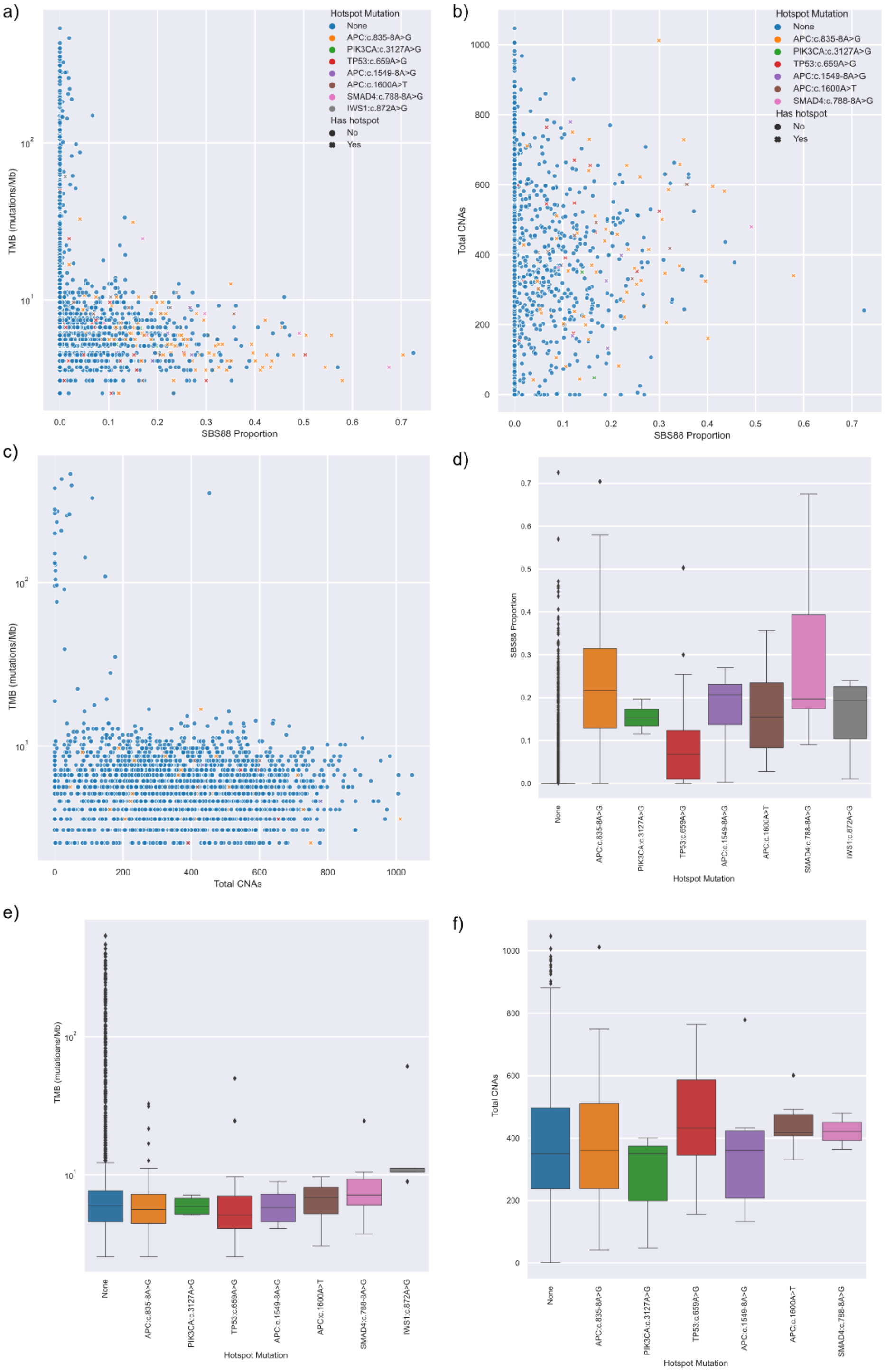


**Supplementary Figure 8:** Relationship between SBS88, TMB, CNA count and somatic hotspot mutation across all analyzed tumors (n=4,308). SBS88 showed an inverse relationship with (a) TMB and (b) CNA count, as well as an increasing proportion of tumors with a hotspot mutation. (c) Only tumors with low CNA count showed ultra-hypermutated TMB. Considering differences between tumors with specific hotspots and other SBS88-positive tumors, *TP53*:c.659A>G showed (d) lower SBS88 proportion on average ($p=3x10^{-9}$) while (e) tumors with *IWS1*:c.872A>G showed higher TMB ($p=3x10^{-18}$).

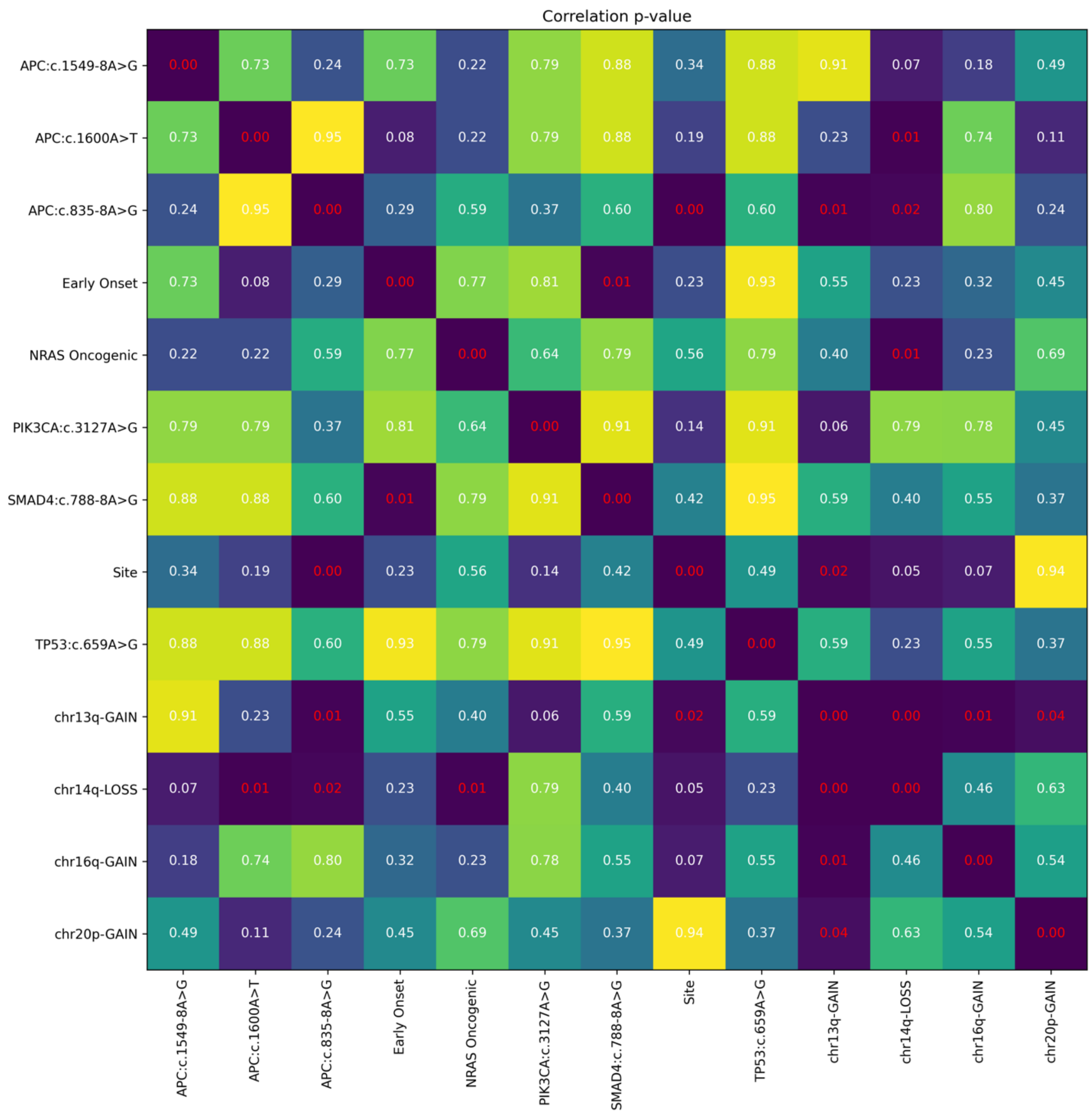


**Supplementary Figure 9**: correlation of covariates in SBS88 positive tumors (limited to those with CNA data). Values shown are p-values, with p-values less than 0.05 highlighted in red.

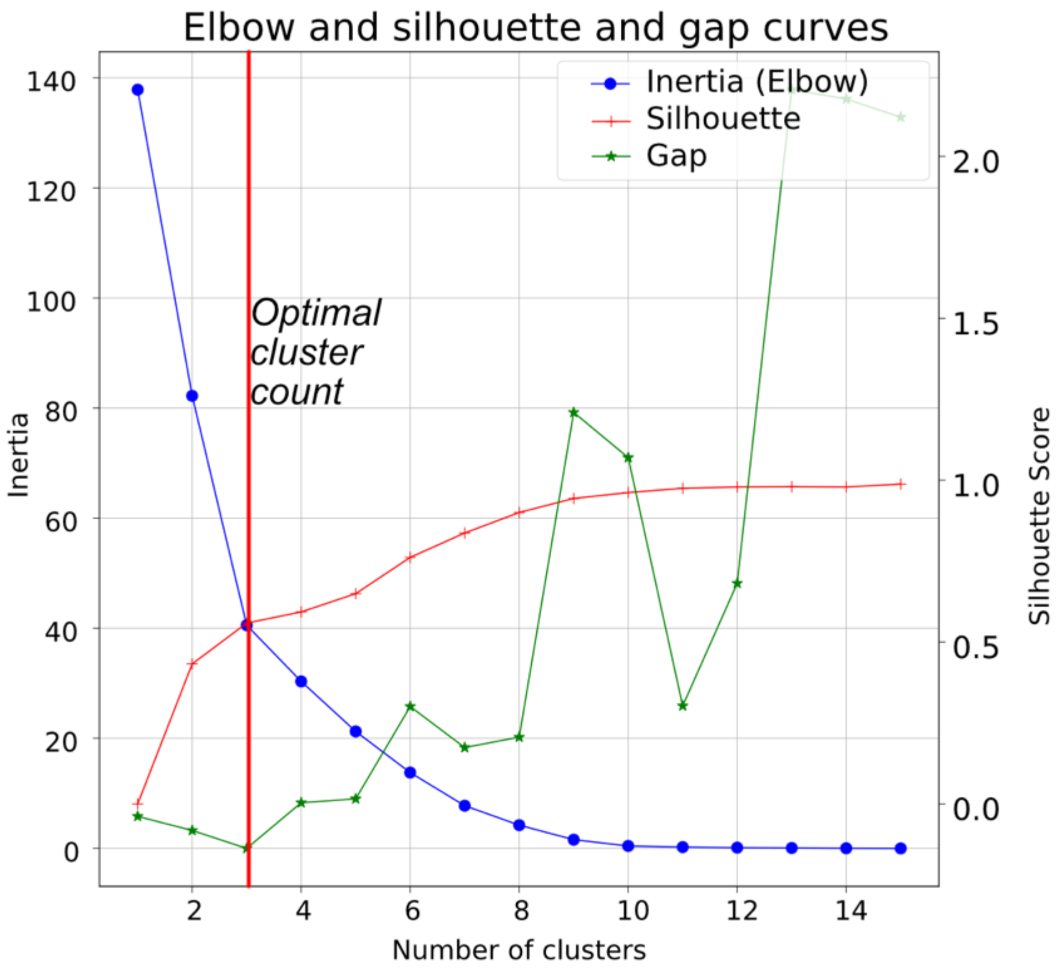


**Supplementary Figure 10**: Three measures were used to determine a likely cluster size of three, then applied to the MCA (multiple correspondence analysis) data to infer cluster membership of each tumor.

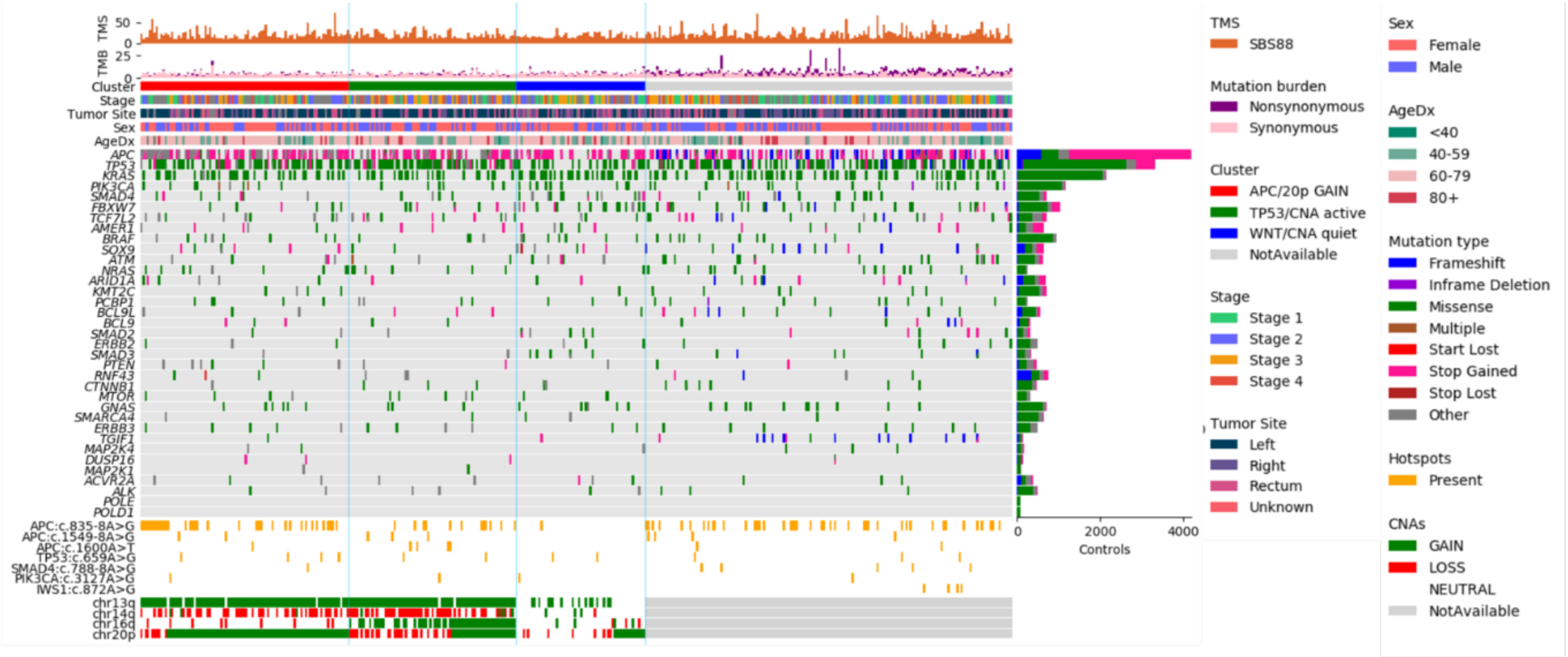


**Supplementary Figure 11:** Oncoplot depicting the distribution of non-silent somatic mutations in CRC-related genes (36) as well as exonuclease domain mutations in *POLE* and *POLD1* in 392 SBS88 positive CRCs, grouped by cluster, and 3,916 SBS88 negative CRCs (controls), as well as significantly associated recurrent hotspot mutations and enriched CNAs.

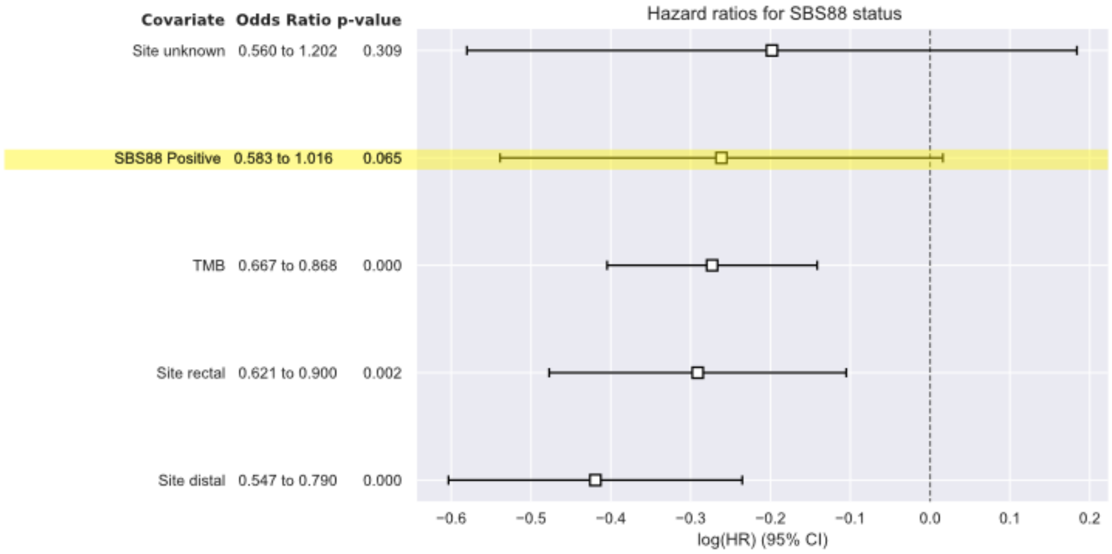


**Supplementary Figure 12:** Survival odds-ratio for SBS88 positive tumors stratified on stage, study, sex and dichotomized age, and adjusting for cancer site and tumor mutational burden (TMB).

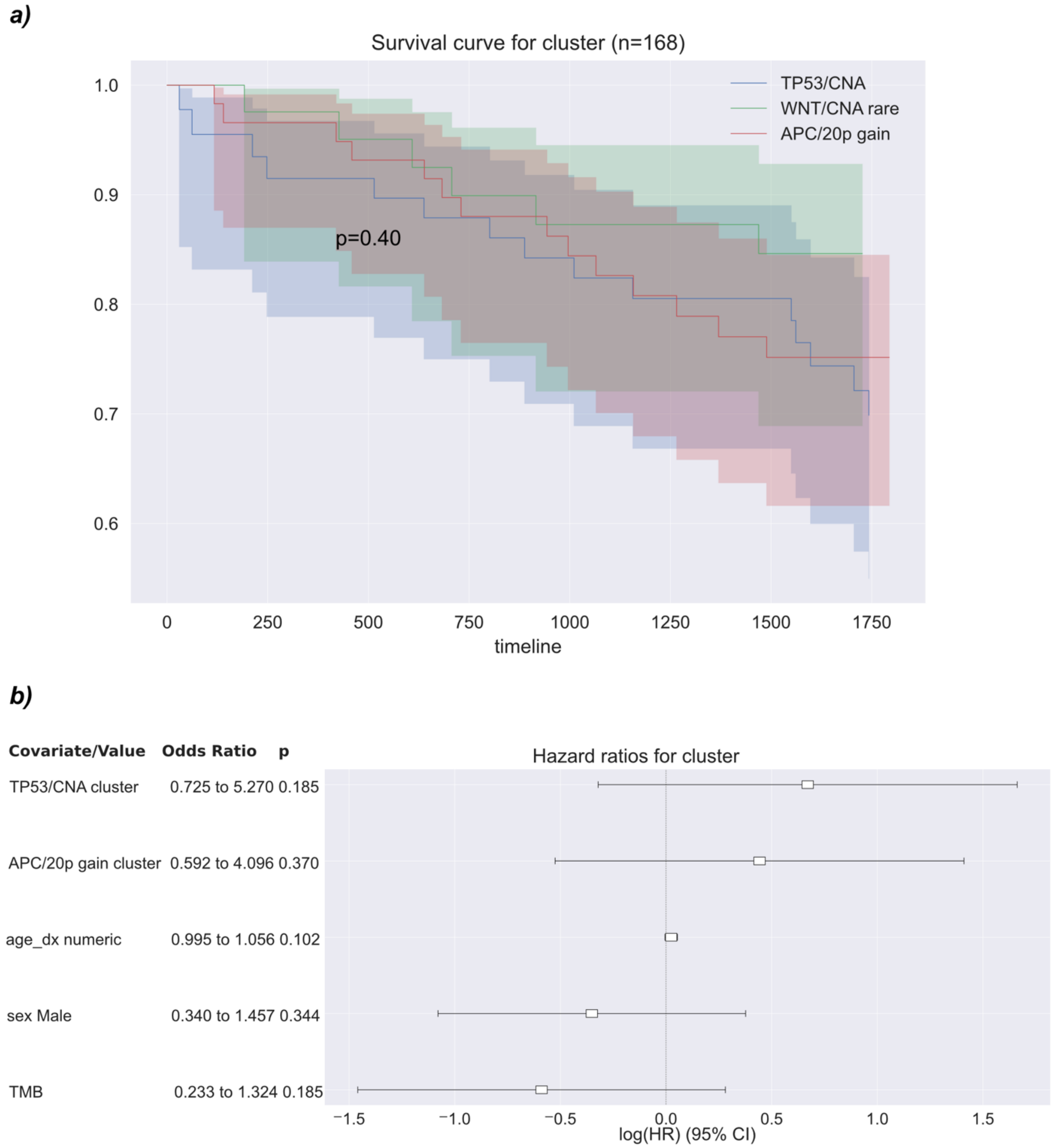


**Supplementary Figure 13**: Survival analysis by cluster did not show significant differences, though the trend suggests cluster 3 (WNT/CNVs rare) has better survival relative to clusters 1 (*APC* hotspots/CNVs) and 2 (*TP53*/CNVs).
